# Social isolation in mid-life: associations with psychological distress, life satisfaction and self-rated health in two successive British birth cohorts

**DOI:** 10.1101/2024.08.05.24311494

**Authors:** Rosie Mansfield, Marcus Richards, George B. Ploubidis, Morag Henderson, Praveetha Patalay

## Abstract

**Purpose:** This study examines how different forms of social isolation, such as living alone, lack of community engagement, and unemployment, are associated with mental health in mid-life (ages 42-46), a life stage often overlooked when examining the impacts of social isolation.

**Methods:** Using longitudinal data (1999-2016) from two British birth cohort studies: 1970 British Cohort Study N=16,585 and the 1958 National Child Development Study N=15,806, this study investigated whether different forms of isolation have independent effects, contribute to cumulative risk, or interact additively or multiplicatively.

**Results:** Independent effects varied by isolation type and mental health outcomes. Being out of employment was linked to higher psychological distress and lower life satisfaction and self-rated health, while living alone was only associated with lower life satisfaction. Limited contact with friends and relatives and a lack of community engagement were associated with lower life satisfaction and self-rated health. Greater social isolation corresponded to increased psychological distress, lower life satisfaction, and poorer self-rated health, demonstrating cumulative risk. Effects appeared additive rather than multiplicative. No consistent sex or cohort differences were observed.

**Conclusion:** The study underscores the need to examine both separate and combined effects of social isolation across the complete mental health state. Isolation in its various forms was detrimental for mental health in mid-life and was most consistently linked to lower life satisfaction. Efforts to reduce isolation and its negative mental health impacts must recognise the complexity of these experiences.

## Introduction

Social isolation is indicated by quantifiable, situational factors across a range of relational contexts such as living alone or infrequent contact with friends, family and people in the community [1]–[3]. It is distinct from, although related to, an individual’s qualitative assessment of the meaning, value and function of social relationships [4], [5], and also how the circumstances arose e.g., chosen solitude. Studies show social isolation is more prevalent than loneliness [6], [7] and independently predicts lower wellbeing and higher mortality in later life [2], [8]–[10]. However, research and policy often conflate the two [11], with few studies focusing primarily on social isolation [12], [13].

Public health researchers are increasingly encouraged to adopt a multi-domain, multi-context approach to studying social isolation [14]. Investigating the effect of individual social isolation experiences across contexts in addition to cumulative risk provides a more nuanced picture, revealing the most toxic social conditions for mental health that are modifiable through policy [3], [5], [6], [14]. For example, although living alone is associated with worse mental health outcomes, it is unlikely a direct risk factor [15]. Frequent contact with friends improves quality of life [16] and social participation, neighbourhood cohesion and physical activity have been shown to attenuate the negative mental health impact of living alone in Japan, Ghana and Finland [17]–[19]. Unemployment also contributes to isolation, carrying long-term mental health consequences and financial barriers to social engagement [20], [21]. Declining community engagement in Britian, worsened by cuts to local services, is also a major concern [22], [23]. Taken together, these findings demonstrate the value of examining social connections across a range of contexts to understand the processes through which isolation and poor mental health are associated such as reduced access to health resources and limited accountability for positive health behaviours [24].

### Social isolation in mid-life

During post-retirement age, people lack work networks, are less likely to have dependent children in the household or within close proximity and are more likely to experience spousal bereavement. It is understandable then, that there is a much larger evidence base relating to social isolation as a risk factor for poor mental health in this population compared to other life stages [25], [26]. There is also a growing body of literature in adolescence showing that positive aspects of social contact are protective factors for mental health [27], [28]. However, the significance of social isolation in mid-life has, until recently, been overlooked, despite being characterised by lower wellbeing [29], [30], and diverse family networks, household compositions, labour market participation, and care responsibilities that bridge the gap between younger and older generations [31]. Social connectedness may therefore be particularly important during mid-life to mitigate against increased life pressures such as career demands and family obligations, and common shifts in family dynamics and roles, such as loss of parents, children leaving home and divorce [32].

Age stratified, cross-sectional analysis of the Swiss Health Survey found a negative association between social isolation and depression across early, mid, and later life [33]. More recently longitudinal analysis using the New Zealand birth cohort provided evidence for accelerated brain ageing and related negative health outcomes in individuals reporting social isolation in mid-life [34], emphasising the importance of this life stage. In the most comprehensive research to date, Luo and Li 2022 studied social isolation trajectories in mid and later life using the United States (US) Health and Retirement Study (HRS) [5]. They identified four patterns of isolation, with the most isolated individuals experiencing worse health outcomes, including functional limitations, depression, memory deficits, and low self-rated health. The healthiest group had the highest social engagement, suggesting social activity may be more beneficial than subjective social support. However, most studies include a social isolation index that combines experiences. This approach does not permit investigations of the independent, interactive, and cumulative associations between specific forms of isolation and mental health, which can reveal modifiable social conditions.

### The current study

Despite increased policy focus on social isolation and loneliness in the UK, there is limited evidence from large scale, population-based studies on social isolation and its relationship to mental health. More research is needed that captures multiple indicators of social isolation across a range of relational contexts, to investigate the independent, cumulative and interactive associations.

Informed by Cornwell and Waite’s (2009) definition of social disconnectedness [1], situational factors that cover an individuals’ social network, infrequent social interactions, and a lack of participation in social activities and groups were captured in the current study. These mapped onto multiple relational contexts within which social isolation can occur (e.g., household, labour market, community) identified in a previous study by the authors to align with Bronfenbrenner’s microsystem (i.e., an individual’ social conditions within their immediate environment) [22]. Multiple outcomes were also included with the aim of covering different dimensional aspects of the complete mental health state including aspects of subjective wellbeing in addition to symptoms of mental-ill health [35]. Given established distinctions between the correlates of mental illness and wellbeing [27], the inclusion of life satisfaction provided the opportunity to examine social isolation beyond its association with mental distress. Similarly, there is increased consensus that self-rated general health is a useful prognostic indicator for depression and is therefore included in the current study as a subjective measure encompassing physical and psychosocial aspects of health [36].

By analysing two successive birth cohorts born in 1958 and 1970, the current study also offers cross-cohort perspectives to better understand if social isolation is associated with mental health consistently over time. It is important to acknowledge that these cohorts are broadly representative of those born in mainland Britain at those times (1958, 1970) and therefore include samples that are predominantly white British. Analyses in the current study were stratified by sex given known differences in social isolation experiences between males and females [22]. For example, males tend to be more socially isolated, as seen in a US study assessing relationships with partners, friends, family, and community [37] and, in England, male isolation remains more stable across the lifecourse than female isolation [38].

Stratification of all analyses by sex, and analysis of cohort effects, provides the opportunity to consider social isolation within different time periods and increasingly diverse and complex lifecourse trajectories with regards to family formation, care, and labour market participation [39], [40]. While gendered patterns are referenced, this study stratifies by sex at birth rather than gender identity.

The aim of the current study was to identify the independent, cumulative and interactive associations of different forms of social isolation with psychological distress, life satisfaction and self-rated general health in mid-life, and to explore sex and cross-cohort differences. We set out to answer the following research questions:

1. What are the independent associations between different forms of social isolation and psychological distress, life satisfaction and self-rated health in mid-life, and are there sex and cohort differences?
2. What is the cumulative association between multiple forms of social isolation and psychological distress, life satisfaction and self-rated health in mid-life, and are there sex and cohort differences?
3. Is the effect of different combinations of social isolation on psychological distress, life satisfaction and self-rated health additive or multiplicative, and are there sex and cohort differences?

## Method

### Data Sources

Data were from two successive British birth cohort studies: the 1970 British Cohort Study (1970 BCS) [41], [42] and the 1958 National Child Development Study (1958 NCDS) [43]. Cohort members were born in Great Britain (i.e., England, Wales & Scotland) in one week of 1970 and 1958 respectively, with regular follow-up surveys from birth. Social isolation was assessed during mid-life (ages 42-46) and data on mental health outcomes was taken at the next available sweep at age 46 (2016) (1970 BCS) and 50 (2008) (1958 NCDS).

### Analytic sample

For both cohorts, the analytic sample was defined by the target population of the most recent sweep used in the analysis i.e., those alive and residing in Great Britian at age 46 in 1970 BCS (N=16,585) and age 50 in 1958 NCDS (N=15,806). This criteria assumed that the mortality rate within cohorts is representative of the population [44]. For a summary of participants’ demographic, socioeconomic, and health characteristics, both imputed and complete case, see Supplementary Tables S1 and S2.

### Measures

#### Social isolation

Social isolation is a multi-dimensional construct measured in this study by a range of self-reported indicators across different relational contexts (e.g., household, community) [22]. Similar items across cohorts were identified indicating social isolation within the household (i.e., living alone), a lack of regular contact with friends and relatives outside of the household, employment status, and a lack of regular community engagement including community groups, religious activity, and volunteering. To ensure completeness of isolation indicators and consistency across cohorts, items were taken from sweeps between the ages of 42-46. For example, in the 1970 BCS, data were from the age 42 sweep except from information relating to cohort members’ engagement with community groups or organisation which was taken at age 46. In the 1958 NCDS, data were from the age 46 sweep with the exception of frequency of contact with friends and relatives outside the household which was captured at age 44. Items were harmonised across cohorts to provide simple indicator variables where 1 = socially isolated e.g., living alone =1, out of employment = 1. The employment indicator was generated by combining information on both education and employment status i.e., out of education and employment, to account for small numbers of cohort members who may have left work to go back into education. For connectedness to family and friends, participants were deemed isolated if they had no regular (at least monthly) contact with friends and relatives outside of the household. An isolation indicator was created for community engagement whereby a participant was deemed isolated if they satisfied two of the three criteria: did not engage in regular (at least monthly) religious activity, were not a volunteer and were not a member of a community group or organisation. To understand the ‘dose-response’ association i.e., the severity of outcomes according to the degree of isolation across contexts, a cumulative social isolation score was also generated. The total score was between 0-4 where 0 = no social isolation and 4 = high social isolation; however, due to small counts for social isolation scores of four, scores of three and four were combined.

Different combinations of indicators were examined using interaction terms to understand if any particular combination of isolation experiences was more strongly associated with mental health in mid-life compared to others. For more detail on the items and coding of social isolation variables, see Supplementary Table S3.

#### Mental Health Outcomes

Psychological distress: In the 1970 BCS at age 46 and in the 1958 NCDS at age 50, psychological distress was captured using the Malaise 9-item Questionnaire [45], [46]. The 9-item version provides a reliable and valid assessment of psychological distress that is consistent within and between generations, suggesting that participants’ understanding of the mental health items is comparable across these two cohorts [47].

Subjective wellbeing – life satisfaction: Subjective wellbeing was captured in both the 1970 BCS and 1958 NCDS using a measure of life satisfaction. Cohort members were asked to indicate on a scale of 0-10 how satisfied or dissatisfied they were with the way their life had turned out so far, where 0 indicates “*completely dissatisfied*” and 10 indicates “*completely satisfied*”.

Self-rated general health: Self-rated general health was measured in both cohorts with the item: ‘In general, would you say your health is…1 “*excellent*” 2 “*very good*” 3 “*good*” 4 “*fair*” 5 “*poor*”?’. The item was recoded so that good self-rated health was indicated by higher scores. Self-rated health is increasingly used as a prognostic indicator for depression. It is therefore included in the current study as a subjective measure capturing the physical and psychosocial aspects of health [36].

Demographic, socioeconomic and health characteristics: Cohort members’ sex was used to stratify analyses to understand possible differences in the impact of social isolation on mental health between males and females. Education (highest level of educational achievement (degree vs. no degree)), was included as a covariate alongside socioeconomic variables: self-reported financial difficulties, occupational social class, and home ownership. Self-rated psychological distress and general health were also included as well as a binary indicator of limiting long-standing illness. Inclusion of a covariate set aimed to minimise possible confounding and socio-economic and health characteristics were taken from the most recent sweep available prior to mid-life in an attempt to avoid any reverse causality. For more detail on the items and coding of demographic, socioeconomic and health covariates, see Supplementary Table S3.

### Analysis Strategy

#### Missing data strategy

To deal with biases in estimates that can arise from non-randomness in discontinued participation and item non-response, multiple imputation (MI) using chained equations was applied separately for each cohort. Based on the overall proportions of missingness in the outcomes and standard recommendations, we chose to run 50 imputations [48], [49] (see Supplementary Table S4. for more information on levels of missing data). Further details on the selection of auxiliary variables are provided in the supplementary materials along with sensitivity analyses using an ‘impute and delete’ method [50] and complete case sample.

#### Independent effects

All analyses were conducted using Stata 17 software [51]. Linear multivariable regression models were run for each separate social isolation indicator with psychological distress, life satisfaction and self-rated general health as the outcomes. This determined the size of effect for different social isolation experiences across contexts. All models were stratified by sex and included the full covariate set and a cohort interaction term estimated to identify differences in association between social isolation and mental health across cohorts. Only the coefficient for the social isolation variable of interest was interpreted in each model to avoid table two fallacy [52].

#### Cumulative effects

The cumulative effect of experiencing multiple forms of social isolation was explored by repeating the above models but replacing the independent social isolation indicator with a cumulative social isolation score between 0-4 where a higher score indicated greater social isolation. Due to low numbers of cohort members reporting four experiences of social isolation, those with a score of three or four were combined to give a scale between 0-3.

#### Additive and multiplicative effects

Two-way interaction terms were created for all six combinations of the four social isolation experiences (e.g., living alone* out of employment), and included in linear multivariable regression models as outlined above. By identifying whether effects were additive or multiplicative, this analysis revealed which combinations of social isolation experiences were most toxic for mental health. Analyses were stratified by sex and an additional three-way interaction term included for social isolation interaction term*cohort to understand any cohort differences. Significant interactions (p<.10) were visualised using margins plots. To explore whether alternative combinations of social isolation experiences existed within the data, a sensitivity latent class analysis was adopted to identify groups of individuals showing qualitatively similar patterns in isolation [53]. More details of the latent class methodology are presented alongside the results in the supplementary file.

## Results

Table 1. presents the descriptive statistics for social isolation indicators and mental health outcomes.

**Table 1.**
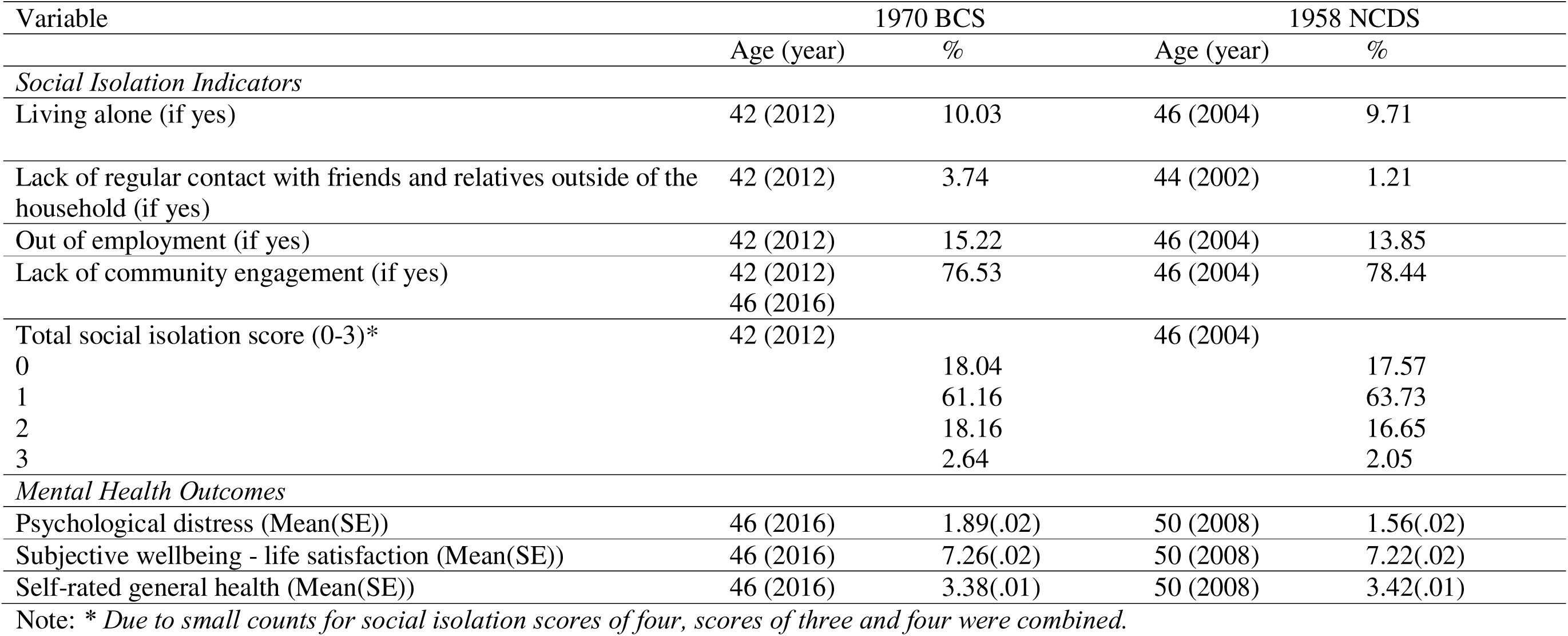
Descriptive statistics for social isolation indicators and mental health outcomes using multiply imputed data m=50 (1970 BCS N = 16,585 and 1958 NCDS = 15,806)

### Independent effects

Figure 1. presents the standardised regression coefficients for the independent associations of social isolation indicators with psychological distress, life satisfaction and self-rated general health. A lack of frequent contact with friends and relatives, no labour market participation and limited community engagement were associated with lower life satisfaction and self-rated general health. However, living alone was only associated with lower life satisfaction. Being out of employment was the social isolation indicator most consistently associated with poorer mental health. Supplementary Table S5. includes all results from the linear multivariable regression models for each separate social isolation indicator to identify independent effects and cohort interaction effects on psychological distress, life satisfaction and self-rated general health using multiply imputed data.

**Figure 1.**
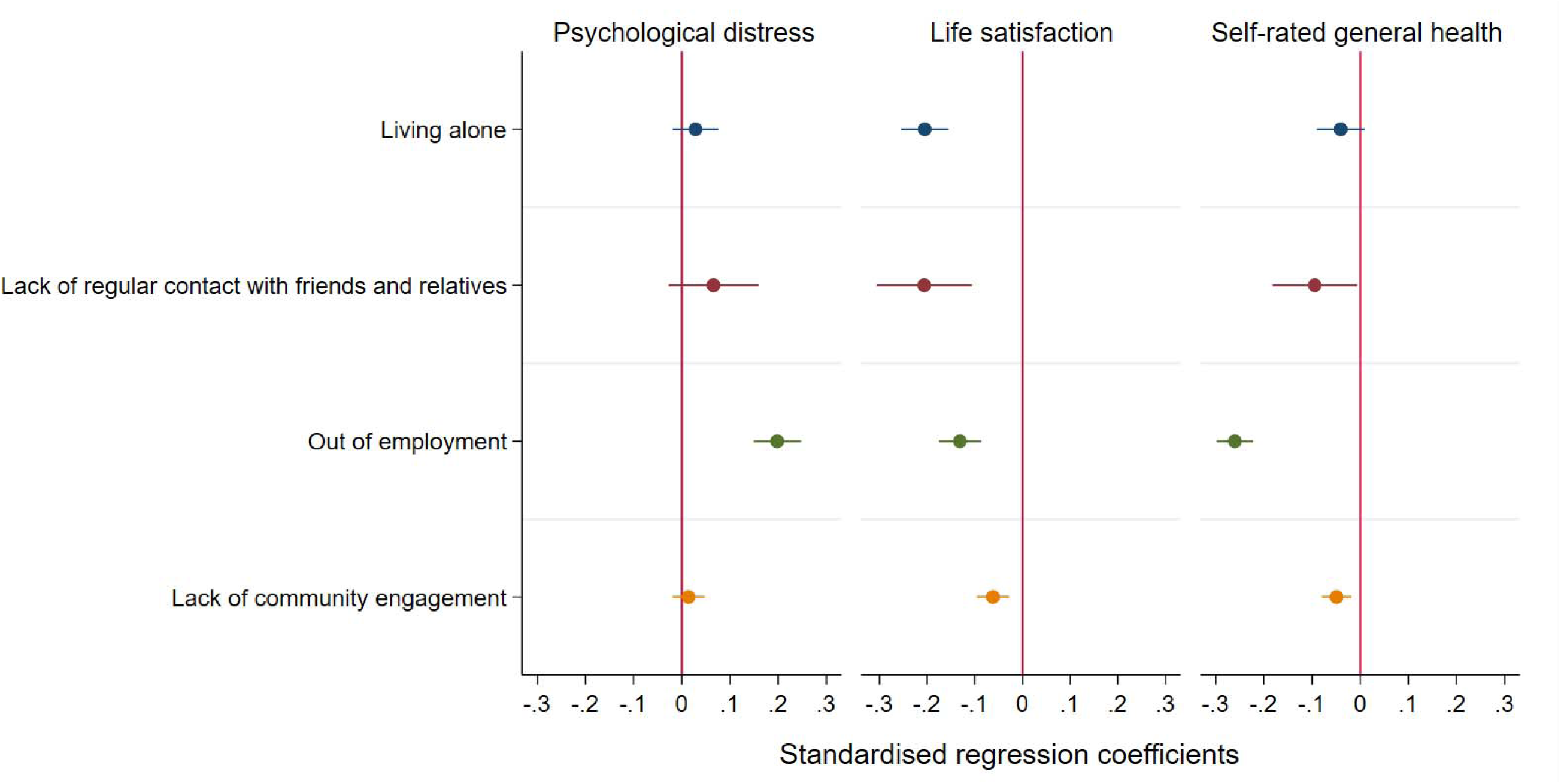
Standardized regression coefficients for the independent associations of social isolation indicators with psychological distress, life satisfaction and self-rated general health (N = 32,391)

The interaction between being out of employment and cohort was associated with life satisfaction and self-rated general health such that, the life satisfaction and general health of those born in 1958 was more negatively impacted by being out of employment when compared with those born in 1970. A similar cohort interaction was present for the association between living alone and self-rated general health such that there was a more negative health impact for those living alone in the 1958 birth cohort. The detrimental effects of some social isolation experiences in mid-life were therefore more pronounced for those born in 1958 (mid-life survey 2004) when compared to those born in 1970 (mid-life survey 2012) (see Supplementary Figure S1. a, b, c and d for marginal mean plots for the significant interaction effects). However, the opposite was found for the association between a lack of frequent contact with friends and relatives and psychological distress, with those born in 1970 experiencing higher psychological distress associated with a lack of frequent contact. No sex differences were found. Models were run with the full set of demographic, socioeconomic and health covariates.

### Cumulative effects

To understand the cumulative effect of experiencing multiple forms of social isolation on mental health outcomes, independent social isolation indicators were replaced with a cumulative social isolation score, where a higher score indicated greater social isolation. Scores ranged from 0-4; however, due to very low frequency of scores of four, top scores were combined resulting in a range of 0-3. Results from the multiply imputed linear multivariable regression models can be found in Table 4. Cumulative social isolation was associated with all mental health outcomes such that the greater the level of social isolation i.e., the more forms of social isolation experienced, the higher the psychological distress and the lower the life satisfaction and self-rated general health. However, results from the independent models indicate that the association between overall level of social isolation and mental health outcomes is driven more by some forms of isolation than by others. The interaction between cumulative social isolation and cohort was associated with self-rated general health. Despite lower levels of self-rated general health for those born in 1970, the difference between the general health of this cohort and those born in 1958 was smallest for cohort members with a high social isolation score of three and much larger for those with low scores of zero. This shows that the self-rated health of the two cohorts is more similar for those with higher levels of social isolation. See Supplementary Figure S2. for the marginal mean plot.

**Table 4.** Linear multivariable regression models for the cumulative social isolation score to identify 1) cumulative effects using the cumulative social isolation score, and 2) cohort effects in any cumulative associations with psychological distress, life satisfaction and self-rated general health –multiply imputed (m=50) models (N = 32,391)

| All coefficients presented are for the<br>cumulative social isolation score as the<br>exposure | Psychological distress |  | Life satisfaction |  | Self-rated general health |  |
| --- | --- | --- | --- | --- | --- | --- |
|  | coef |  | coef |  | coef |  |
|  | [95% CI] |  | [95% CI] |  | [95% CI] |  |
|  | N |  | N |  | N |  |
|  | (1)Cumulative<br>effect | (2)*Cohort | (1)Cumulative<br>effect | (2)*Cohort | (1)Cumulative<br>effect | (2)*Cohort |
| *Full sample model* | .070***<br>[.046 - .094]<br>32,391 | -.007<br>[-.045 - .032]<br>32,391 | -.119***<br>[-.143 - -.094]<br>32,391 | -.003<br>[-.040 - .034]<br>32,391 | -.106***<br>[-.127 - -.086]<br>32,391 | -.081***<br>[-.122 - -.039]<br>32,391 |
| *Males only* | .064***<br>[.033 - .095]<br>15,810 | -.011<br>[-.061 - .039]<br>15,810 | -.128***<br>[-.159 - -.097]<br>15,810 | -.006<br>[-.057 - .046]<br>15,810 | -.108***<br>[-.137 - -.079]<br>15,810 | -.087***<br>[-.144 - -.031]<br>15,810 |
| *Females only* | .079***<br>[.050 - .109]<br>15,221 | -.009<br>[-.061 - .044]<br>15,221 | -.108***<br>[-.139 - -.077]<br>15,221 | -.000<br>[-.052 - .051]<br>15,221 | -.108***<br>[-.132 - -.083]<br>15,221 | -.071***<br>[-.122 - -.020]<br>15,221 |

Note: All models are adjusted for the full covariate set and are run once with a cohort dummy variable and again to include a cohort interaction term (\*cohort). Coefficients are reported for cumulative and cohort interaction effects.
\*\*\* $p < 0.01$ , \*\* $p < 0.05$ , \* $p < 0.1$

### Additive and multiplicative effects

The only significant interactions (p<.10) were being out of employment by living alone on life satisfaction and being out of employment by lack of community engagement on self-rated general health. Cohort members out of employment had lower life satisfaction and general health overall; however, the difference in scores between those in and out of employment was greatest for those living alone and lacking community engagement. Individuals who lived alone and were out of employment reported the lowest levels of life satisfaction.

Furthermore, those lacking community engagement showed poorer self-rated general health compared with cohort members engaging more with their community. This difference was much larger for individuals who were also out of employment. These interactions were found to be significant in the same direction in the female only sample but not in the male only sample (see Figure 2.). No additional three-way interactions by cohort were observed. For the full set of results from the linear multivariable regression models using multiply imputed data, see Supplementary Table S9.

**Figure 2.**
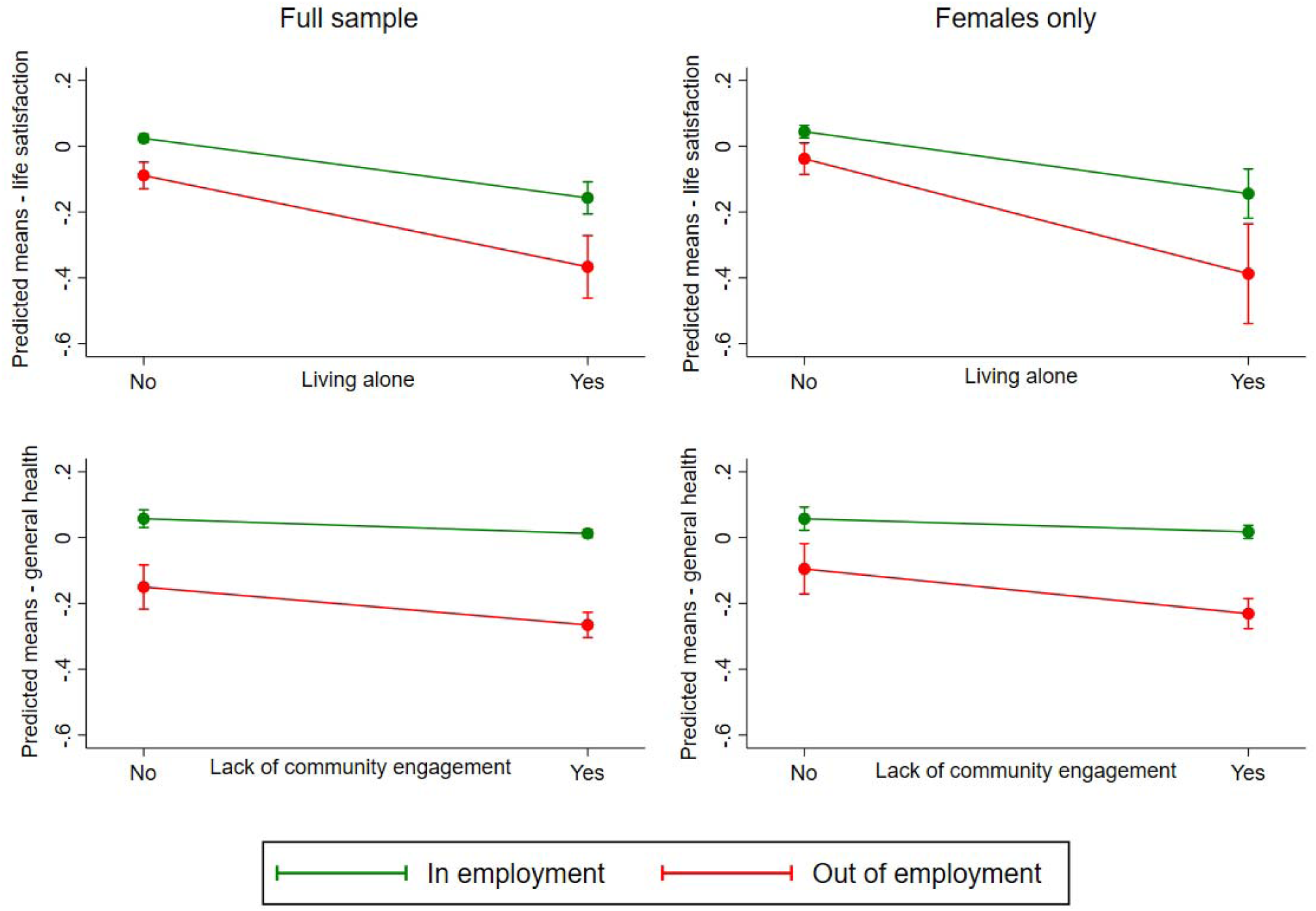
Model predicted marginal mean plots for significant (p<.10) interactions between multiple social isolation indicators in the full imputed sample (N = 32,391) and for females only (N = 15,221)

### Sensitivity analyses

Results from independent effects models were checked against models including all social isolation indicators (i.e., effects net of other isolation experiences), yielding similar conclusions. Complete case and impute-and-delete models were also comparable (see Supplementary Tables S6–S8, S10, S11). Latent class models tested one to five classes to identify alternative social isolation patterns. A three-class solution was optimal (Supplementary Table S12), with males and the 1970 BCS cohort more likely in isolated classes (Supplementary Table S13). However, these classes did not differ qualitatively from the two-way interaction models, so class membership was not analysed against mental health outcomes.

## Discussion

The current study captured multiple indicators of social isolation to investigate the independent, cumulative and interactive associations with psychological distress, life satisfaction and self-rated general health in mid-life. Stratifying analyses by sex and cohort (1970 BCS, 1958 NCDS) allowed for examining social isolation across different time periods and evolving lifecourse trajectories in family, care, and labour market participation [31].

After controlling for demographic, socioeconomic, and health factors, infrequent contact with friends and relatives and limited community engagement were associated with lower life satisfaction and poorer self-rated health but not higher psychological distress, aligning with research on older adults [16]. In this study, infrequent contact with friends and relatives (less than monthly) was uncommon, whereas limited community engagement, measured by lack of group membership, religious activity and volunteering, was widespread [22]. Regular community engagement may boost wellbeing for a minority, but a significant increase in psychological distress is not observed perhaps due to its absence being normalised. In the UK, cuts to local government funding for community and cultural services over the past 15 years have worsened health and well-being [23]. Limited membership of clubs and organisations will likely be driven by a decline in the availability of these community groups and services and also individuals’ resources. Findings highlight the need to reinvest in community assets, supported by research showing the long-term benefits of social participation for physical activity in mid-life [54].

Research shows higher psychological distress and lower life satisfaction among those living alone [15], but in this study, living alone was *only* associated with lower life satisfaction. This highlights the relevance of investigating social isolation in relation to the complete mental health state including subjective wellbeing [35]. Life satisfaction, a global quality-of-life measure, reflects overall wellbeing, while psychological distress refers to impairing symptoms of emotional suffering such as anxiety and depression. Though overlapping, mental illness and well-being have distinct correlates, supporting their separation in research to avoid conflation [27]. Overall, in this study, social isolation was most strongly and consistently linked to lower life satisfaction.

Being out of employment in mid-life was associated with poorer mental health across all outcomes, supporting a recent US study that found disconnection from work in early adulthood was a risk factor for depressive symptoms in mid-life [20]. While being out of employment had negative mental health impacts for both sexes, males appeared more vulnerable, possibly due to traditional gender roles and reliance on economic success [21]. The financial strain associated with being out of employment, which includes having to engage with welfare systems, further impacts wellbeing [55]. However, the current study cannot determine the extent to which negative associations between being out of employment and mental health were due to a lack of social contact or other psychological and financial aspects related to work that are known to affect wellbeing [21]. Similarly, despite investigating social isolation in mid-life and mental health outcomes a few years later, this study does not account for health problems or caring responsibilities in mid-life that may be, for example, the reason for unemployment.

In addition to findings relating to independent indicators of social isolation, higher cumulative social isolation scores were associated with greater psychological distress, lower life satisfaction, and poorer self-rated general health aligning with existing literature on mid [5], [32]–[34] and older adults [2], [8]–[10]. However, given that independent associations varied by form of isolation and specific mental health outcome, results indicate that the association between overall level of social isolation and mental health could mask important discrepancies between rates and mental health impacts of different isolation experiences. This has implications for producing policies that can reduce specific forms of isolation most toxic for mental health [5], [14], and is support for future research that comprehensively investigates multiple forms of isolation across contexts and both their independent and combined effects [6], [56].

The detrimental mental health effects of some social isolation experiences in mid-life were more pronounced for those born in 1958 (mid-life survey in 2004) when compared to those born in 1970 (mid-life survey in 2012). For example, those born in 1958 were more negatively impacted by living alone and being out of employment, with lower life satisfaction and self-rated general health. Despite greater economic precarity in 2012, unemployment had a weaker association with mental health in the 1970 cohort, possibly because joblessness was more common, reducing stigma [21].While the 1970 cohort had lower overall self-rated health when compared to those born in 1958, highly isolated individuals in both cohorts showed similarly poor health, indicating that social isolation has remained consistently negative for general health over time.

Analysis showed little evidence that different forms of isolation amplify each other’s negative mental health effects, suggesting an additive rather than multiplicative impact. Two exceptions were observed whereby cohort members lacking community engagement had poorer self-rated health, especially if out of employment, and those living alone reported lower life satisfaction, particularly if also unemployed. These results were found only in the female sample, which is important given we know that women’s labour market trajectories are complex [39], with more time out in mid-life [22]. Policies should prioritise flexible labour market options for parents and enhance community involvement opportunities for women out of employment during mid-life to support their mental health.

Males had a greater likelihood of membership to the more isolated classes, and the association between living alone and poorer self-rated general health was also approaching significance in the male sample with no association found for females. These findings support previous studies that suggest men are more isolated across the lifecourse than females [37], [38] and could indicate that men living alone have reduced positive health behaviours [24]. Study members born in the 1970 BCS cohort were also more likely to belong to classes experiencing multiple forms of isolation, supporting previous research that showed an increase in some forms of social isolation over time in the UK [22].

This study provides evidence from two large scale, population-based studies on social isolation in mid-life and its relationship to mental health in Great Britian. Longitudinal cohort studies enable researchers to account for reverse causality by controlling for baseline mental health and establishing a clear temporal order between social isolation and mental health [57]. However, this study, like many other observational studies, will have some reverse causation bias and there is the possibility of confounding due to related factors [58].

Investigating multiple indicators of social isolation across a range of relational contexts enabled a better understanding of the specific conditions leading to poor mental health and the stability of these associations over time. Despite including multiple social isolation indicators, the current study was limited by the information available across both studies. More comprehensive measurement of social networks and frequency of contact, for example, number of friends, may have provided the opportunity to model unique clusters of social isolation experiences against the various outcomes. To ensure completeness of isolation indicators and consistency across cohorts, items were taken from sweeps between the ages of 42-46 with outcomes at the next available sweep four years later. Differences in the timing of social isolation exposures across cohorts may explain a proportion of the cohort effects.

However, cohort differences were not seen across all models, indicating that the year of data collection did not have universal effects. All analysed data were collected prior to the Covid-19 pandemic. Evidence from the British cohort studies reveals large inequalities in experiences of social isolation and loneliness in older adults prior to Covid-19 and that the pandemic worsened the extent of these [7]. We might therefore expect that levels of isolation in mid-life have increased beyond what is reported in this study.

The cohorts used in the current study are broadly representative of those born in mainland Britain at those times (1958, 1970) and therefore include samples that are predominantly white British. Therefore, the generalisability of results to non-white British populations is limited. As mid-life data becomes available in young and more diverse British birth cohorts, the ethnic differences in the association between social isolation and mental health should be explored.

## Conclusion

Evidence of cumulative risk was found with higher social isolation scores associated with greater psychological distress, lower life satisfaction, and poorer self-reported general health. Independent associations varied by form of isolation, justifying future research that investigates both the individual and combined effects of different social isolation experiences. In the current study, the effects of different combinations of social isolation on mental health appeared to be additive. Findings varied by outcome, with stronger and more consistent associations between social isolation and lower life satisfaction when compared with psychological distress and self-rated general health.

## Data Availability

Data openly available via the UK Data Service:
University College London, UCL Social Research Institute, Centre for Longitudinal Studies. (2024). 1970 British Cohort Study. [data series]. 11th Release. UK Data Service. SN: 200001, DOI: http://doi.org/10.5255/UKDA-Series-200001 University College London, UCL Social Research Institute, Centre for Longitudinal Studies. (2024). National Child Development Study. [data series]. 14th Release. UK Data Service. SN: 2000032, DOI: http://doi.org/10.5255/UKDA-Series-2000032

## Acknowledgements

We are extremely grateful to cohort members of the two cohorts included in this research for their participation over the years. We thank our collaborators at the What Works Centre for Wellbeing and our Scientific Advisory Group for their engagement and input.

## References

[1] Cornwell EY, Waite LJ (2009) Measuring social isolation among older adults using multiple indicators from the NSHAP study. Journals Gerontol - Ser B Psychol Sci Soc Sci 64(SUPPL.1):38–46. 10.1093/geronb/gbp037

[2] Holt-Lunstad J, Smith TB, Baker M, Harris T, Stephenson D (2015) Loneliness and social isolation as risk factors for mortality: A meta-analytic review. Perspect Psychol Sci 10(2):227–237. 10.1177/1745691614568352

[3] Huisman M, van Tilburg TG (2021) Social exclusion and social isolation in later life. In: Ferraro KF, Carr D (ed) Handbook of Aging and the Social Sciences, 9th edn. Elsivier Academic Press, pp 99–114.

[4] Hughes JT, Waite LJ, Hawkley LC, Cacioppo JT (2004) A short scale for measuring loneliness in large surveys: Results from two population-based studies. Res Aging 26(6):655–672. 10.1109/ICIT.2017.7913066

[5] Luo M, Li L (2022) Social isolation trajectories in midlife and later-life: patterns and associations with health. Int J Geriatr Psychiatry 37(5). 10.1002/gps.5715

6. [6] d’Hombres B, Barjaková M, Schnepf SV (2021) Loneliness and social isolation: An unequally shared burden in Europe. No. 14245, p. 23. Available: https://www.iza.org/publications/dp/14245/loneliness-and-social-isolation-an-unequally-shared-burden-in-europe

[7] Mansfield R et al. (2023) Examining the inter-relationships between social isolation and loneliness and their correlates among older British adults before and during the COVID-19 lockdown: Evidence from four British longitudinal studies. Innov Aging, 8(1), igad126, 10.1093/geroni/igad126

[8] Coyle CE, Dugan E (2012) Social isolation, loneliness and health among older adults. J Aging Health 24(8):1346–1363. 10.1177/0898264312460275

[9] Golden J et al. (2009) Loneliness, social support networks, mood and wellbeing in community-dwelling elderly. Int J Geriatr Psychiatry 24(7):694–700. 10.1002/gps.2181

[10] Steptoe A, Shankar A, Demakakos P, Wardle J (2013) Social isolation, loneliness, and all-cause mortality in older men and women. Proc Natl Acad Sci USA 110(15):5797– 5801. 10.1073/pnas.1219686110

[11] Wigfield A (2022) Developing a new conceptual framework of meaningful interaction for understanding social isolation and loneliness. Soc Policy Soc 21(2):172–193

[12] Holt-Lunstad J, Steptoe A (2022) Social isolation: An underappreciated determinant of physical health. Curr Opin Psychol 43:232–237. 10.1016/j.copsyc.2021.07.012

[13] Loades ME et al. (2020) Rapid systematic review: The impact of social isolation and loneliness on the mental health of children and adolescents in the context of COVID-19. J Am Acad Child Adolesc Psychiatry 59(11):1218–1239.e3. 10.1016/j.jaac.2020.05.009

[14] Klinenberg E (2016) Social isolation, loneliness, and living alone: Identifying the risks for public health. Am J Public Health 106(5):786–787. 10.2105/AJPH.2016.303166

[15] McElroy E et al. (2023) Living alone and mental health: Parallel analyses in UK longitudinal population surveys and electronic health records prior to and during the COVID-19 pandemic. BMJ Ment Health 26(1):1–10. 10.1136/bmjment-2023-300842

[16] Luna E et al. (2020) The prospective association between frequency of contact with friends and relatives and quality of life in older adults from Central and Eastern Europe. Soc Psychiatry Psychiatr Epidemiol 55(8):1001–1010. 10.1007/s00127-020-01834-8

[17] Kondo K, Kawachi I, Kondo N (2018) Older adults in Japan: The JAGES longitudinal study. 28(7):2–7

[18] Gyasi RM, Abass K, Adu-Gyamfi S (2020) How do lifestyle choices affect the link between living alone and psychological distress in older age? Results from the AgeHeaPsyWel-HeaSeeB study. BMC Public Health 20(1):1–9. 10.1186/s12889-020-08870-8

[19] Pasanen TP, Tamminen N, Martelin T, Solin P (2021) Positive mental health of Finnish people living alone: The role of circumstantial factors and leisure-time activities. Int J Environ Res Public Health 18(13). 10.3390/ijerph18136735

[20] Caniglia M (2024) Nonparticipation in work and education in emerging adulthood and depressive symptoms through early midlife. SSM - Ment Health 6(June):100329. 10.1016/j.ssmmh.2024.100329

[21] Virgolino A et al. (2022) Lost in transition: A systematic review of the association between unemployment and mental health. J Ment Health 31(3):432–444. 10.1080/09638237.2021.2022615

[22] Mansfield R, Henderson M, Richards M, Ploubidis GB, Patalay P (2024) Lifecourse trajectories and cross-generational trends in social isolation: Findings from five successive British birth cohort studies. Adv Life Course Res 60(October 2023):100613. 10.1016/j.alcr.2024.100613

[23] Fahy K et al. (2023) Mental health impact of cuts to local government spending on cultural, environmental and planning services in England: A longitudinal ecological study. BMC Public Health 23(1):1–13. 10.1186/s12889-023-16340-0

[24] Berkman LF (2008) Social work in health care: Social support, social networks, social cohesion and health. Behav Soc Work Health Care Settings 1389(June):37–41. 10.1300/J010v31n02

[25] Smith KJ, Victor C (2019) Typologies of loneliness, living alone and social isolation, and their associations with physical and mental health. Ageing Soc 39(8):1709–1730. 10.1017/S0144686X18000132

[26] Blazer D (2012) Social isolation and loneliness in older adults—A mental health/public health challenge. Int J Geriatr Psychiatry 27(7): 739–749. 10.1002/gps.3799

[27] Patalay P, Fitzsimons E (2016) Correlates of mental illness and wellbeing in children: Are they the same? Results from the UK Millennium Cohort Study. J Am Acad Child Adolesc Psychiatry 55(9): 771–783. 10.1016/j.jaac.2016.05.019

[28] Christiansen J, Larsen FB, Lasgaard M (2021) Associations of loneliness and social isolation with physical and mental health among adolescents and young adults. Perspect Public Health 141(4): 226–236. 10.1177/17579139211016077

[29] Blanchflower DG (2021) Is happiness U-shaped everywhere? Age and subjective well-being in 145 countries. J Popul Econ 34(2): 575–624. 10.1007/s00148-020-00797-z

[30] Gondek D, Moltrecht B, Ploubidis G (2021) Mental health crisis in midlife–a proposed research agenda. Res Ideas Outcomes 7:e62024. 10.3897/rio.7.e62024

[31] Lachman ME (2015) Mind the gap in the middle: A call to study midlife. Res Hum Dev 12(3–4): 327–334. 10.1080/15427609.2015.1068048

[32] Sehmi R, Maughan B, Matthews T, Arseneault L (2020) No man is an island: Social resources, stress, and mental health at mid-life. Br J Psychiatry 217(5): 638–644. 10.1192/bjp.2019.25

[33] Hämmig O (2019) Health risks associated with social isolation in general and in young, middle, and old age. PLoS One 14(7): e0219663. 10.1371/journal.pone.0219663

[34] Lay-Yee R, Hariri AR, Knodt AR, Barrett-Young A, Matthews T, Milne BJ (2023) Social isolation from childhood to mid-adulthood: Is there an association with older brain age? Psychol Med. 10.1017/S0033291723001964

35. [35] World Health Organization (2022) Transforming mental health for all. Available: https://www.who.int/publications/i/item/9789240049338

[36] Ambresin G, Chondros P, Dowrick C, Herrman H, Gunn JM (2014) Self-rated health and long-term prognosis of depression. Ann Fam Med 12(1): 57–65. 10.1370/afm.1562

[37] Umberson D, Lin Z, Cha H (2022) Gender and social isolation across the life course. J Health Soc Behav. 10.1177/00221465221109634

[38] Read S, Comas-Herrera A, Grundy E (2020) Social isolation and memory decline in later life. J Gerontol B Psychol Sci Soc Sci 75(2): 367–376. 10.1093/geronb/gbz152

[39] Widmer ED, Ritschard G (2009) The de-standardization of the life course: Are men and women equal? Adv Life Course Res 14(1–2): 28–39. 10.1016/j.alcr.2009.04.001

[40] Roberson PNE, Norona JC, Zorotovich J, Dirnberger Z (2017) Developmental trajectories and health outcomes among emerging adult women and men. Emerg Adulthood 5(2): 128–142. 10.1177/2167696816662118

[41] Elliott J, Shepherd P (2006) Cohort profile: 1970 British Birth Cohort (BCS70). Int J Epidemiol 35(4): 836–843. 10.1093/ije/dyl174

[42] Sullivan A, Brown M, Hamer M, Ploubidis GB (2023) Cohort profile update: The 1970 British Cohort Study (BCS70). Int J Epidemiol 52(3): e179–e186. 10.1093/ije/dyac148

[43] Power C, Elliott J (2006) Cohort profile: 1958 British birth cohort (National Child Development Study). Int J Epidemiol 35(1): 34–41. 10.1093/ije/dyi183

[44] Mostafa T, Gambaro L, Sironi M, Goodman A, Silverwood RJ, Ploubidis GB (2021) Missing at random assumption made more plausible: Evidence from the 1958 British birth cohort. J Clin Epidemiol 136: 44–54. 10.1016/j.jclinepi.2021.02.019

[45] Rodgers B, Pickles A, Power C, Collishaw S, Maughan B (1999) Validity of the Malaise Inventory in general population samples. Soc Psychiatry Psychiatr Epidemiol 34(6): 333–341. 10.1007/s001270050153

[46] Rutter WK, Tizard J (1970) Education, health and behaviour. London: Longmans.

[47] Ploubidis GB, McElroy E, Moreira HC (2019) A longitudinal examination of the measurement equivalence of mental health assessments in two British birth cohorts. Longitud Life Course Stud 10(4):471–489.

[48] Sullivan A, Brown M, Hamer M, Ploubidis GB (2023) Cohort Profile Update: The 1970 British Cohort Study (BCS70). Int J Epidemiol 1–8. 10.1093/ije/dyac148

[49] Lee JH, Huber JC (2021) Evaluation of multiple imputation with large proportions of missing data: How much is too much? Iran J Public Health 50(7):1372–1380.

50. Von Hippel PT (2007) Regression with missing Ys: An improved strategy for analyzing multiply imputed data. Sociol. Methodol. 10.1111/j.1467-9531.2007.00180.x

51. StataCorp (2021) Stata Statistical Software: Release 17.

[52] Westreich D, Greenland S (2013) The table 2 fallacy: Presenting and interpreting confounder and modifier coefficients. Am J Epidemiol 177(4):292–298. 10.1093/aje/kws412

[53] Weller BE, Bowen NK, Faubert SJ (2020) Latent Class Analysis: A Guide to Best Practice. J Black Psychol 46(4):287–311. 10.1177/0095798420930932

[54] Tsoli S, Fancourt D, Sullivan A, Hamer M, Ploubidis GB, Kawachi I (2024) Life-course social participation and physical activity in midlife: longitudinal associations in the 1970 British Cohort Study (BCS70). Eur J Epidemiol 39(6):643–651. 10.1007/s10654-024-01107-7

[55] Wickham S, Bentley L, Rose T, Whitehead M, Taylor-Robinson D, Barr B (2020) Effects on mental health of a UK welfare reform, Universal Credit: a longitudinal controlled study. Lancet Public Heal 5(3):e157–e164. 10.1016/S2468-2667(20)30026-8

[56] Huisman M, van Tilburg TG (2021) Social exclusion and social isolation in later life. In: Ferraro KF, Carr D (ed) Handbook of Aging and the Social Sciences, 9th edn. Elsivier Academic Press, pp 99–114.

[57] Seifert N, Seddig D, Eckhard J (2022) Does social isolation affect physical and mental health? A test of the social causation hypothesis using dynamic panel models with fixed effects. Aging Ment Heal 26(7):1353–1367. 10.1080/13607863.2021.1961125

[58] Liu B, Floud S (2017) Unravelling the associations between social isolation, loneliness, and mortality. Lancet Public Heal 2(6):e248–e249. 10.1016/S2468-2667(17)30090-7

