## Supplementary File S1 for "Social isolation in mid-life: associations with psychological distress, life satisfaction and self-rated health in two successive British birth cohorts" (from another version of this preprint: version 2, 2024-12-02)

**Social isolation in mid-life: associations with psychological distress, life satisfaction and self-rated health in two successive British birth cohorts**

Rosie Mansfield [a*] Marcus Richards [b], George B. Ploubidis [a], Morag Henderson [a+], Praveetha Patalay [a, b+]

[a] Centre for Longitudinal Studies, University College London

[b] MRC Unit for Lifelong Health and Ageing, University College London

+Shared senior authors

Figure S2. *Model predicted marginal mean plot for cumulative social isolation score by cohort interaction on self-rated general health* ... 25

Table S8. *Linear multivariable regression models for the cumulative social isolation score to identify 1) 'dose response' effects using the cumulative social isolation score, and 2) cohort effects in any 'dose-response' associations on psychological distress, life satisfaction and self-rated general health – complete case and impute and delete models* ... 26

Table S9. *Linear multivariable regression models for the interaction between multiple social isolation indicators to identify 1) the additive and multiplicative interaction effects, and 2) cohort effects in any interaction associations on psychological distress, life satisfaction and self-rated general health using multiply imputed data m=50 (Full sample N = 32,391, Males N = 15,810, Females N = 15,221)* ... 28

Table S10. *Linear multivariable regression models for the interaction between multiple social isolation indicators to identify 1) the additive and multiplicative interaction effects, and 2) cohort effects in any interaction associations on psychological distress, life satisfaction and self-rated general health using complete case data* ... 30

Table S11. *Linear multivariable regression models for the interaction between multiple social isolation indicators to identify 1) the additive and multiplicative interaction effects, and 2) cohort effects in any interaction associations on psychological distress, life satisfaction and self-rated general health using impute and delete data* ... 33

*Latent Class Analysis Methodology* ... 35

Table S12. *Results from the latent class analyses (1-5 classes) including goodness-of-fit statistics (full sample with 1970 BCS and 1958 cohorts combined N=19,902)* ... 35

Table S13. *Class membership (N(%)) for full sample and by sex and cohort* ... 36

Table S1. *Descriptive statistics for demographic, socioeconomic and health covariates, social isolation indicators and mental health outcomes using complete case data*

| | 1970 BCS | | | 1958 NCDS | | |
|---|---|---|---|---|---|---|
| Variable | N | Age(year) | % | N | Age(year) | % |
| *Demographic, Socioeconomic and Health Covariates* | | | | | | |
| Sex (if female) | 15,226 | Birth | 7,457(48.98) | 15,805 | Birth | 7,764(49.12) |
| Self-reported sex (if female) | 8,804 | 38 (2008) | 4,638(52.68) | 11,213 | 42 (1999) | 5,712(50.94) |
| Education – highest qualification - educated to degree level (if no degree) | 8,764 | 38 (2008) | 8,418(96.05) | 15,805 | 42 (1999) | 15,325(96.96) |
| Socioeconomic status – self-reported financial difficulties (if yes) | 8,773 | 38 (2008) | 797(9.08) | 11,174 | 42 (1999) | 896(8.02) |
| Socioeconomic status – occupational social class | 7,423 | 38 (2008) | | 9,487 | 42 (1999) | |
| Professional/Managerial-technical | | | 3,739(50.37) | | | 4,059(42.78) |
| Intermediate | | | 2,771(37.33) | | | 3,937(41.50) |
| Partly skilled/Unskilled | | | 881(11.87) | | | 1,471(15.51) |
| Others | | | 32(0.43) | | | 20(0.21) |
| Socioeconomic status – homeownership (if no) | 8,757 | 38 (2008) | 1,922(21.95) | 11,131 | 42 (1999) | 2,091(18.79) |
| Health – self-rated general health (Mean (SE)) | 8,770 | 38 (2008) | 3.73(1.01) | 11,171 | 42 (1999) | 3.09(0.76) |
| Health – limiting long standing illness (if yes) | 8,803 | 38 (2008) | 1,394(15.84) | 11,171 | 42 (1999) | 3,234(28.95) |
| Health – psychological distress (Mean (SE)) | 9,445 | 34 (2004) | 1.67(1.90) | 11,076 | 42 (1999) | 1.51(1.78) |
| Health – life satisfaction (Mean (SE)) | 9,441 | 34 (2004) | 7.41(1.80) | 11,069 | 42 (1999) | 7.30(1.91) |
| *Social Isolation Indicators* | | | | | | |
| Living alone (if yes) | 9,802 | 42 (2012) | 949(9.68) | 9,432 | 46 (2004) | 870(9.22) |
| Lack of regular contact with friends and relatives outside of the household (if yes) | 9,749 | 42 (2012) | 345(3.54) | 8,482 | 44 (2002) | 79(0.93) |
| Out of employment (if yes) | 9,767 | 42 (2012) | 1,400(14.33) | 9,448 | 46 (2004) | 1,164(12.32) |
| Lack of community engagement (if yes) | 6,512 | 42 (2012)<br>46 (2016) | 4,869(74.77) | 9,441 | 46 (2004) | 7,259(76.89) |

| | | | | | | |
|---|---|---|---|---|---|---|
| Total social isolation score (0-3)*<br>0<br>1<br>2<br>3 | 6,497 | 42 (2012) | <br>1,307(20.12)<br>4,062(62.52)<br>1,022(15.73)<br>106(1.63) | 7,816 | 46 (2004) | <br>1,546(19.78)<br>5,078(64.97)<br>1,088(13.92)<br>104(1.33) |
| *Mental Health Outcomes* | | | | | | |
| Psychological distress (Mean(SD)) | 7,866 | 46 (2016) | 1.77(2.13) | 7,866 | 50 (2008) | 1.77(2.13) |
| Subjective wellbeing - life satisfaction (Mean(SD)) | 8,487 | 46 (2016) | 7.35(1.89) | 9,631 | 50 (2008) | 7.29(1.84) |
| Self-rated general health (Mean(SD)) | 8,576 | 46 (2016) | 3.44(1.10) | 9,733 | 50 (2008) | 3.48(1.11) |

Note: * Due to small counts for social isolation scores of four, scores of three and four are combined.

Table S2. *Descriptive statistics for demographic, socioeconomic and health covariates using multiply imputed data m=50 (1970 BCS N = 16,585 and 1958 NCDS = 15,806)*

| | 1970 BCS | | 1958 NCDS | |
|---|---|---|---|---|
| Variable | Age (year) | % | Age (year) | % |
| Self-reported sex (if female) | 38 (2008) | 53.17 | 42 (1999) | 50.64 |
| Education – highest qualification - educated to degree level (if no degree) | 38 (2008) | 96.18 | 42 (1999) | 96.96 |
| Socioeconomic status – self-reported financial difficulties (if yes) | 38 (2008) | 10.13 | 42 (1999) | 8.40 |
| Socioeconomic status – occupational social class | 38 (2008) | | 42 (1999) | |
| Professional/Managerial-technical | | 45.96 | | 38.31 |
| Intermediate | | 38.88 | | 42.44 |
| Partly skilled/Unskilled | | 14.51 | | 18.92 |
| Others | | 0.66 | | 0.33 |
| Socioeconomic status – homeownership (if no) | 38 (2008) | 24.65 | 42 (1999) | 20.10 |
| Health – self-rated general health (Mean(SE)) | 38 (2008) | 3.67(.01) | 42 (1999) | 3.07(.01) |
| Health – limiting long standing illness (if yes) | 38 (2008) | 17.18 | 42 (1999) | 29.81 |
| Health – psychological distress (Mean(SE)) | 34 (2004) | 1.75(.02) | 42 (1999) | 1.55(.02) |
| Health – life satisfaction (Mean(SE)) | 34 (2004) | 7.33(.02) | 42 (1999) | 7.27(.02) |

Table S3. *Harmonisation of demographic, socioeconomic and health covariates, social isolation indicators and mental health outcomes*

| Measure | Cohort | Age (year) | Variable | Item/Questionnaire | Recode |
|---|---|---|---|---|---|
| *Demographic, Socioeconomic and Health Covariates* | | | | | |
| Sex | 1970 BCS | 38 (2008) | b8cmsex | Self-reported cohort member's sex | Males = 0 Females = 1 recode 1=0 2=1 |
| | 1958 NCDS | 42 (1999) | n622_6 | Self-reported cohort member's sex | Males = 0 Females = 1 recode 1=0 2=1 |
| Education – highest qualification – educated to degree level | 1970 BCS | 38 (2008) | bd8achq1 | Highest Academic Qualification CM obtained in 2008 survey<br>-1 Level not calculable<br>0 No academic qualification<br>1 GCSE D-E<br>2 Other Scottish Quals<br>3 GCSE A-C, Scot Standards/Intermediate 1<br>4 Scot Intermediate 2 / AS Levels or 1 A Level<br>5 2+ A Levels, Scot Higher/6th<br>6 Diploma<br>7 Degree, PGCE, Other Degree qual<br>8 Higher Degree | Recode Degree, PGCE, Other Degree qual<br>8 Higher Degree = 0 anything else = 0 and no degree = 1 |
| | 1958 NCDS | 42 (1999) | numdeg | No. of degrees CM has obtained? | Recode any degree = 0 no degree = 1 |

| | | | | | |
|---|---|---|---|---|---|
| Socioeconomic status – self-reported financial difficulties | 1970 BCS | 38 (2008) | b8finnow | How well would you say you personally are managing financially these days. Would you say you are<br>1 ...living comfortably<br>2 doing all right<br>3 just about getting by<br>4 finding it quite difficult or<br>5 finding it very difficult? | Recode 1 living comfortably 2 doing all right and 3 just about getting by = 0 and 4 finding it quite difficult and 5 finding it very difficult = 1 |
| | 1958 NCDS | 42 (1999) | finnow | How well would you say you personally are managing financially these days. Would you say you are<br>1 ...living comfortably<br>2 doing all right<br>3 just about getting by<br>4 finding it quite difficult or<br>5 finding it very difficult? | Recode 1 living comfortably 2 doing all right and 3 just about getting by = 0 and 4 finding it quite difficult and 5 finding it very difficult = 1 |
| Socioeconomic status – occupational social class | 1970 BCS | 38 (2008) | b8sc | (Current Job) Social Class<br>1 I Professional<br>2 II Managerial-technical<br>3.1<br>3.2<br>4 IV Partly skilled<br>5 Unskilled<br>6 Others | Recode<br>I Professional<br>II Managerial-technical = 0<br>3.1/3.2 = 1<br>IV Partly skilled<br>Unskilled = 2<br>Others = 3 |
| | 1958 NCDS | 42 (1999) | sc | (Current Job) Social Class<br>1 I Professional<br>2 II Managerial-technical | Recode<br>I Professional |

| | | | | | |
|---|---|---|---|---|---|
| | | | | 3.1<br>3.2<br>4 IV Partly skilled<br>5 Unskilled<br>6 Others | II Managerial-technical = 0<br>3.1/3.2 = 1<br>IV Partly skilled<br>Unskilled = 2<br>Others = 3 |
| Socioeconomic status – home ownership | 1970 BCS | 38 (2008) | b8ten2 | Do you own or rent your home or have some other arrangement?<br>1 Own - outright<br>2 Own - buying with help of a mortgage<br>3 Pay part rent and part mortgage<br>4 Rent it<br>5 Live here rent-free<br>6 Squatting<br>7 Other | Recode<br>1 Own - outright<br>2 Own - buying with help of a mortgage<br>3 Pay part rent and part mortgage = 0<br>4 Rent it<br>5 Live here rent-free<br>6 Squatting<br>7 Other = 1 |
| | 1958 NCDS | 42 (1999) | tenure2 | Do you own or rent your home or have some other arrangement?<br>1 Own - outright<br>2 Own - buying with help of a mortgage<br>3 Pay part rent and part mortgage<br>4 Rent it<br>5 Live here rent-free<br>6 Squatting<br>7 Other | Recode<br>1 Own - outright<br>2 Own - buying with help of a mortgage<br>3 Pay part rent and part mortgage = 0<br>4 Rent it<br>5 Live here rent-free<br>6 Squatting<br>7 Other = 1 |

| | | | | | |
|---|---|---|---|---|---|
| Health – self-rated general health | 1970 BCS | 38 (2008) | b8hlthgn | In general, would you say your health is...<br>1 Excellent<br>2 Very good<br>3 Good<br>4 Fair<br>5 Poor | Recode so that high scores = better health 5 = "Excellent", 1 = "Poor" |
| | 1958 NCDS | 42 (1999) | hlthgen | How would you describe your health generally? Would you say it is ...<br>1 Excellent<br>2 Good<br>3 Fair<br>4 Poor | Recode so that high scores = better health 4 = "Excellent", 1 = "Poor" |
| Health – limiting long standing illness | 1970 BCS | 38 (2008) | b8lsiotr<br>b8khllt | Whether CM Has Any (Other) Long Standing Illness/ Disability<br>Whether Health Limits Everyday Activities | Combine items to get complete picture – if limiting long standing illness = 1 |
| | 1958 NCDS | 42 (1999) | lsiany2 | Do you have any long-standing illness, disability or infirmity? By long-standing I mean anything that has troubled you over a period of time, or that is likely to affect you over a period of time? Yes/no | Recode yes = 1 no = 0 |
| Health – psychological distress | 1970 BCS | 34 (2004) | b7mal02<br>b7mal03<br>b7mal05<br>b7mal09<br>b7mal12<br>b7mal14<br>b7mal16 | Malaise 9-item Questionnaire | Total score |

| | | | | | |
|---|---|---|---|---|---|
| | | | b7mal20 b7mal21 | | |
| | 1958 NCDS | 42 (1999) | mal02 mal03 mal05 mal09 mal12 mal14 mal16 mal20 mal21 | Malaise 24-item Questionnaire reduced down to match the 9-item Questionnaire | Total score |
| Health – life satisfaction | 1970 BCS | 34 (2004) | b7lifet1 | Here is a scale from 0 to 10, where '0' means that you are completely dissatisfied and '10' means that you are completely satisfied. Please enter the number which corresponds with how satisfied or dissatisfied you are with the way life has turned out so far | 0-10 |
| | 1958 NCDS | 42 (1999) | lifesat1 | Here is a scale from 0-10 where '0' means that you are completely dissatisfied and '10' means that you are completely satisfied. Please enter the number which corresponds with how satisfied or dissatisfied you are about the way your life has turned out so far | 0-10 |
| *Social Isolation Indicators* | | | | | |
| Living alone | 1970 BCS | 42 (2012) | BD9HSIZE B9GSLIVE B9GRTOK | Derived Household Size (bcs70_2012_derived.dta) HHgrid: whether person living with CM at interview (bcs70_2012_persongrid.dta) HHgrid: Person's relationship to CM (bcs70_2012_persongrid.dta) | Derived variable for living alone where living alone = 1 using household grid and household member's relationship to cohort member – |

| | | | | | |
|---|---|---|---|---|---|
| | | | | | cross-checked against the study derived variable for household size |
| | 1958 NCDS | 46 (2004) | nd7numhh<br>n7rtok12-n7rtok20 | (Derived) Total number of people in household<br>Person's relationship to Cohort Member | Derived variable for living alone where living alone = 1 using household grid and household member’s relationship to cohort member – cross-checked against the study derived variable for household size |
| Lack of regular contact with friends and relatives outside of the household | 1970 BCS | 42 (2012) | B9FREMT<br>B9FAMMT | How often do you meet up with any of your friends? (bcs70_2012_flatfile.dta)<br>How often do you meet up with any members of your family who you do not live with? (bcs70_2012_flatfile.dta) | Recode to combine frequency of contact with family and friends and to indicate social isolation i.e., less than monthly contact = 1 |
| | 1958 NCDS | 44 (2002) | seerels<br>visrels<br>numpals | How often do you have regular contact with relatives outside your household? | Recode to combine frequency of |

| | | | | | |
|---|---|---|---|---|---|
| | | | | How often do you visit or are you visited by relatives who live outside your household?<br>How often do you have regular contact with friends or acquaintances outside your household? | contact with family and friends and to indicate social isolation i.e., less than monthly contact = 1 |
| Out of employment | 1970 BCS | 42 (2012) | BD9ECACT | (Derived) Current economic activity status | Recode so that being in neither education nor employment = 1 |
| | 1958 NCDS | 46 (2004) | nd7ecact | (Derived) Current economic activity status | Recode so that being in neither education nor employment = 1 |
| Lack of community engagement | 1970 BCS | 42 (2012)<br>46 (2016) | B10Q26<br>B10Q27<br>B9SCQ23<br>B9SCQ1W<br>B9SCQ1X | Whether CM is currently a member of any organisations, clubs or societies? How often CM participates in meetings/events/activities of any organisations?<br>How often, if ever, do you attend any kind of religious service or meeting?<br>How often have you done each of the following activities in the last 12 months? (voluntary organisation) (voluntary work) | Generate variables for membership to community group or organisation, volunteering and regular religious activity (i.e., at least monthly) then generate social isolation indicator for community context - isolated across 2/3 no group |

| | | | | | |
|---|---|---|---|---|---|
| | | | | | membership, no volunteering and no regular religious activity = 1 |
| | 1958 NCDS | 46 (2004) | nd7hobby<br>nd7othg<br>n7rnoweq<br>nd7youth<br>nd7polit<br>nd7eco<br>nd7othv<br>nd7local | I'd like you to think about any groups, clubs or organisations that you've been involved with since we last saw you in [^date of last interview]<br>How often, if ever, do you attend any kind of religious service or meeting? | Generate variables for membership to community group or organisation, volunteering and regular religious activity (i.e., at least monthly) then generate social isolation indicator for community context - isolated across 2/3 no group membership, no volunteering and no regular religious activity = 1 |
| Total social isolation score | 1970 BCS | 42 (2012) | Living alone<br><br>Lack of regular contact with | Sum of all social isolation indicators – capped at 3 i.e., those scoring 4 combined with those scoring 3 due to very low counts. | Sum of all social isolation indicators (0-3) |
| | 1958 NCDS | 46 (2004) | | | |

| | | | | | |
|---|---|---|---|---|---|
| | | | friends and relatives outside of the household<br><br>Out of employment<br><br>Lack of community engagement | | |
| *Mental Health Outcomes* | | | | | |
| Psychological distress | 1970 BCS | 46 (2016) | B10Q28A<br>B10Q28B<br>B10Q28C<br>B10Q28D<br>B10Q28E<br>B10Q28F<br>B10Q28G<br>B10Q28H<br>B10Q28I | Malaise 9-item Questionnaire | Total score |
| | 1958 NCDS | 50 (2008) | N8MAL02<br>N8MAL03<br>N8MAL05<br>N8MAL09<br>N8MAL12<br>N8MAL14<br>N8MAL16<br>N8MAL20<br>N8MAL21 | Malaise 9-item Questionnaire | Total score |

| | | | | | |
|---|---|---|---|---|---|
| Subjective wellbeing - life satisfaction | 1970 BCS | 46 (2016) | B10LIFST1 | Here is a scale from 0 to 10, where '0' means that you are completely dissatisfied and '10' means that you are completely satisfied. Please enter the number which corresponds with how satisfied or dissatisfied you are with the way life has turned out so far | 0-10 |
| | 1958 NCDS | 50 (2008) | N8LIFET1 | Here is a scale from 0 to 10, where '0' means that you are completely dissatisfied and '10' means that you are completely satisfied. Please enter the number which corresponds with how satisfied or dissatisfied you are with the way life has turned out so far | 0-10 |
| Self-rated general health | 1970 BCS | 46 (2016) | B10HLTHGN | In general, would you say your health is 1. excellent 2. very good 3. good 4. fair 5. poor? | Reverse code so that excellent health = 5 |
| | 1958 NCDS | 50 (2008) | N8HLTHGN | In general, would you say your health is 1. excellent 2. very good 3. good 4. fair 5. poor? | Reverse code so that excellent health = 5 |

Table S4. *N(%) of missing cases for covariates, social isolation indicators and mental health outcomes (Full sample N = 32,391, 1970 BCS N = 16,585, 1958 NCDS N = 15,806)*

| Measure | Cohort | Age (year) | N(%) missing | Study outcome non-response |
|---|---|---|---|---|
| *Demographic, Socioeconomic and Health Covariates* | | | | |
| Sex (at birth)<br>Self-reported sex | 1970 BCS | 38 (2008) | 1,359(8.19)<br>7,781(46.92) | - |
| | 1958 NCDS | 42 (1999) | 1(0.01)<br>4,593(29.06) | - |
| Education – highest qualification - educated to degree level | 1970 BCS | 38 (2008) | 7,821(47.16) | - |
| | 1958 NCDS | 42 (1999) | 1(0.01) | - |
| Socioeconomic status – self-reported financial difficulties | 1970 BCS | 38 (2008) | 7,812(47.10) | - |
| | 1958 NCDS | 42 (1999) | 4,632(29.31) | - |
| Socioeconomic status – occupational social class | 1970 BCS | 38 (2008) | 9,162(55.24) | - |
| | 1958 NCDS | 42 (1999) | 6,319(39.98) | - |
| Socioeconomic status – home ownership | 1970 BCS | 38 (2008) | 7,828(47.20) | - |
| | 1958 NCDS | 42 (1999) | 4,675(29.58) | - |
| Health – self-rated general health | 1970 BCS | 38 (2008) | 7,815(47.12) | - |
| | 1958 NCDS | 42 (1999) | 4,635(29.32) | - |
| Health – limiting long standing illness | 1970 BCS | 38 (2008) | 7,782(46.92) | - |
| | 1958 NCDS | 42 (1999) | 4,635(29.32) | - |
| Health – psychological distress | 1970 BCS | 34 (2004) | 7,140(43.05) | - |
| | 1958 NCDS | 42 (1999) | 4,730(29.93) | - |
| Health – life satisfaction | 1970 BCS | 34 (2004) | 7,144(43.08) | - |
| | 1958 NCDS | 42 (1999) | 4,737(29.97) | |

| | | | | |
|---|---|---|---|---|
| *Social Isolation Indicators* | | | | |
| Living alone | 1970 BCS | 42 (2012) | 6,783(40.90) | - |
| | 1958 NCDS | 46 (2004) | 6,374(40.33) | - |
| Lack of regular contact with friends and relatives outside of the household | 1970 BCS | 42 (2012) | 6,836(41.22) | - |
| | 1958 NCDS | 44 (2002) | 7,324(46.34) | - |
| Out of employment | 1970 BCS | 42 (2012) | 6,818(41.11) | - |
| | 1958 NCDS | 46 (2004) | 6,358(40.23) | - |
| Lack of community engagement | 1970 BCS | 42 (2012)<br>46 (2016) | 10,073(60.74) | - |
| | 1958 NCDS | 46 (2004) | 6,365(40.27) | - |
| Total social isolation score (0-3)* | 1970 BCS | 42 (2012) | 10,088(60.83) | - |
| | 1958 NCDS | 46 (2004) | 7,990(50.55) | - |
| *Mental Health Outcomes* | | | | |
| Psychological distress | 1970 BCS | 46 (2016) | 8,719(52.57) | 715(4.31%) |
| | 1958 NCDS | 50 (2008) | 6,172(39.05) | 156(0.99%) |
| Subjective wellbeing - life satisfaction | 1970 BCS | 46 (2016) | 8,098(48.83) | 94(0.57%) |
| | 1958 NCDS | 50 (2008) | 6,175(39.07) | 159(1.01%) |
| Self-rated general health | 1970 BCS | 46 (2016) | 8,009(48.29) | 5(0.03%) |
| | 1958 NCDS | 50 (2008) | 6,073(38.42) | 57(0.36%) |

Note: *8,004(48.26%) of cases in 1970 BCS were unproductive in the most recent data collection sweep (age 46) i.e., they did not take part at all. Any missingness above and beyond 8,004 cases in the outcome is due to item non-response. 6,016(38.06%) unproductive cases in 1958 NCDS. * Due to small counts for social isolation scores of four, scores of three and four are combined.*

Table S5. *Linear multivariable regression models for each separate social isolation indicator to identify (1) independent effects of each social isolation indicator and (2) cohort effects in these independent effects on psychological distress, life satisfaction and self-rated general health using multiply imputed data m=50 (Full sample N = 32,391, Males N = 15,810, Females N = 15,221)*

| All coefficients presented are for each independent social isolation indicator only (exposure) | Psychological distress coef [95% CI] | | Life satisfaction coef [95% CI] | | Self-rated general health coef [95% CI] | |
|---|---|---|---|---|---|---|
| | (1)Independent | (2)*Cohort | (1)Independent | (2)*Cohort | (1)Independent | (2)*Cohort |
| *Living alone* | | | | | | |
| *Full sample model* | .029 [-.019 - .076] | .008 [-.081 - .097] | -.205*** [-.254 - -.155] | .069 [-.026 - .164] | -.040 [-.090 - .009] | -.139*** [-.231 - -.047] |
| *Males only* | .026 [-.031 - .083] | -.016 [-.124 - .091] | -.203*** [-.262 - -.144] | .082 [-.033 - .197] | -.053* [-.114 - .008] | -.133** [-.248 - -.018] |
| *Females only* | .020 [-.044 - .085] | .048 [-.078 - .174] | -.223*** [-.291 - -.155] | .052 [-.092 - .196] | -.038 [-.104 - .029] | -.137** [-.262 - -.012] |
| *Lack of regular contact with friends and relatives outside of the household* | | | | | | |
| *Full sample model* | .066 [-.027 - .159] | -.190* [-.406 - .027] | -.206*** [-.306 - -.106] | .078 [-.145 - .301] | -.094** [-.182 - -.006] | -.062 [-.284 - .160] |
| *Males only* | .042 [-.074 - .158] | -.191 [-.449 - .068] | -.198*** [-.314 - -.083] | .051 [-.209 - .310] | -.091* [-.199 - .018] | -.058 [-.336 - .220] |
| *Females only* | .081 [-.043 - .205] | -.204 [-.523 - .116] | -.232*** [-.375 - -.088] | .098 [-.257 - .453] | -.095 [-.222 - .033] | -.081 [-.385 - .223] |
| *Out of employment* | | | | | | |
| *Full sample model* | .198*** [.149 - .248] | -.027 [-.117 - .062] | -.131*** [-.176 - -.086] | -.080** [-.149 - -.010] | -.260*** [-.298 - -.222] | -.186*** [-.266 - -.106] |
| *Males only* | .252*** [.181 - .323] | -.029 [-.149 - .090] | -.147*** [-.215 - -.078] | -.093* [-.200 - .014] | -.305*** [-.366 - -.245] | -.238*** [-.357 - -.118] |
| *Females only* | .183*** [.128 - .238] | -.046 [-.150 - .058] | -.096*** [-.149 - -.043] | -.071 [-.160 - .019] | -.222*** [-.267 - -.177] | -.157*** [-.248 - -.066] |
| *Lack of community engagement* | | | | | | |

| | | | | | | |
|---|---|---|---|---|---|---|
| *Full sample model* | .014<br>[-.019 - .048] | .021<br>[-.041 - .083] | -.062***<br>[-.096 - -.028] | -.007<br>[-.072 - .057] | -.049***<br>[-.079 - -.019] | -.002<br>[-.066 - .062] |
| *Males only* | -.002<br>[-.047 - .042] | .028<br>[-.054 - .111] | -.074***<br>[-.117 - -.030] | -.032<br>[-.119 - .055] | -.051**<br>[-.093 -.010] | -.010<br>[-.097 - .077] |
| *Females only* | .024<br>[-.018 - .065] | .017<br>[-.068 - .102] | -.055**<br>[-.100 - -.009] | .013<br>[-.069 - .095] | -.049**<br>[-.087 - -.010] | .011<br>[-.069 - .092] |

Note: *All models use multiply imputed data, adjusted for the full covariate set and are run once with a cohort dummy variable and again to include a cohort interaction term (*cohort). Mental health outcomes are standardised, and continuous covariates centred. Coefficients are reported for independent and cohort interaction effects. Male and female samples do not = total sample due to some missing data on the sex at birth variable. Additional models were run for each social isolation indicator controlling for all other indicators to assess the independent associations net of the other social isolation domains – results were consistent with the above models.*

**** p<0.01, ** p<0.05, * p<0.1*

Table S6. *Linear multivariable regression models for each separate social isolation indicator to identify (1) independent effects of each social isolation indicator and (2) cohort effects in these independent effects on psychological distress, life satisfaction and self-rated general health – complete case*

| All coefficients presented are for each independent social isolation indicator only (exposure) | Psychological distress<br>coef<br>[95% CI]<br>*N* | | Life satisfaction<br>coef<br>[95% CI]<br>*N* | | Self-rated general health<br>coef<br>[95% CI]<br>*N* | |
|---|---|---|---|---|---|---|
| | (1)Independent | (2)*Cohort | (1)Independent | (2)*Cohort | (1)Independent | (2)*Cohort |
| *Living alone* | | | | | | |
| *Full sample model* | .011<br>[-.040 - .062]<br>*11,806* | .041<br>[-.060 - .142]<br>*11,806* | -.177***<br>[-.231 - -.124]<br>*12,114* | .035<br>[-.071 - .142]<br>*12,114* | -.066**<br>[-.118 - -.014]<br>*12,160* | -.138***<br>[-.242 - -.035]<br>*12,160* |
| *Males only* | -.002<br>[-.063 - .059]<br>*5,822* | -.017<br>[-.140 - .106]<br>*5,822* | -.175***<br>[-.243 - -.106]<br>*5,972* | .055<br>[-.082 - .192]<br>*5,972* | -.080**<br>[-.148 - -.012]<br>*6,000* | -.129*<br>[-.265 - .007]<br>*6,000* |
| *Females only* | .051<br>[-.037 - .139]<br>*5,663* | .143<br>[-.033 - .320]<br>*5,663* | -.188***<br>[-.277 - -.099]<br>*5,794* | .000<br>[-.177 - .177]<br>*5,794* | -.061<br>[-.145 - .023]<br>*5,811* | -.116<br>[-.284 - .052]<br>*5,811* |
| *Lack of regular contact with friends and relatives outside of the household* | | | | | | |
| *Full sample model* | .029<br>[-.079 - .137]<br>*11,256* | -.226*<br>[-.467 - .015]<br>*11,256* | -.211***<br>[-.323 - -.099]<br>*11,562* | -.024<br>[-.279 - .230]<br>11,562 | -.031<br>[-.140 - .078]<br>*11,604* | -.182<br>[-.430 - .066]<br>*11,604* |
| *Males only* | .075<br>[-.055 - .205]<br>*5,531* | -.331**<br>[-.609 - -.054]<br>*5,531* | -.226***<br>[-.367 - -.085]<br>*5,681* | -.062<br>[-.369 - .245]<br>*5,681* | -.085<br>[-.226 - .055]<br>*5,710* | -.094<br>[-.401 - .212]<br>*5,710* |
| *Females only* | -.025<br>[-.211 - .160]<br>*5,404* | -.089<br>[-.532 - .353]<br>*5,404* | -.152<br>[-.338 - .035]<br>*5,533* | -.014<br>[-.463 - .435]<br>*5,533* | .065<br>[-.113 - .243]<br>*5,545* | -.335<br>[-.763 - .094]<br>*5,545* |

| | | | | | | |
|---|---|---|---|---|---|---|
| *Out of employment* | | | | | | |
| *Full sample model* | .238***<br>[.170 - .305]<br>*11,807* | .090<br>[-.048 - .227]<br>*11,807* | -.198***<br>[-.269 - -.127]<br>*12,114* | -.154**<br>[-.297 - -.010]<br>*12,114* | -.302***<br>[-.371 - -.234]<br>*12,160* | -.147**<br>[-.286 - -.009]<br>*12,160* |
| *Males only* | .290***<br>[.178 - .402]<br>*5,824* | .111<br>[-.127 - .348]<br>*5,824* | -.245***<br>[-.370 - -.120]<br>*5,974* | -.412***<br>[-.676 - -.148]<br>*5,974* | -.442***<br>[-.565 - -.320]<br>*6,002* | -.454***<br>[-.713 - -.195]<br>*6,002* |
| *Females only* | *.228****<br>*[.139 - .317]*<br>*5,662* | .034<br>[-.147 - .216]<br>*5,662* | -.159***<br>[-.249 - -.070]<br>*5,792* | -.066<br>[-.247 - .115]<br>*5,792* | -.236***<br>[-.320 - -.151]<br>*5,809* | -.010<br>[-.181 - .161]<br>*5,809* |
| *Lack of community engagement* | | | | | | |
| *Full sample model* | .021<br>[-.011 - .054]<br>*11,288* | -.023<br>[-.089 - .043]<br>*11,288* | -.065***<br>[-.100 - -.030]<br>*11,364* | -.009<br>[-.080 - .062]<br>*11,364* | -.032*<br>[-.066 - .002]<br>*11,403* | .001<br>[-.067 - .069]<br>*11,403* |
| *Males only* | -.001<br>[-.044 - .042]<br>*5,565* | -.001<br>[-.090 - .087]<br>*5,565* | -.087***<br>[-.137 - -.038]<br>*5,597* | -.033<br>[-.133 - .067]<br>*5,597* | -.062**<br>[-.110 - -.013]<br>*5,622* | -.002<br>[-.101 - .097]<br>*5,622* |
| *Females only* | .044*<br>[-.006 - .094]<br>*5,440* | -.033<br>[-.135 - .068]<br>*5,440* | -.040<br>[-.091 - .011]<br>*5,481* | -.001<br>[-.104 - .102]<br>*5,481* | -.012<br>[-.060 - .037]<br>*5,495* | .031<br>[-.067 - .129]<br>*5,495* |

Note: *All models use complete case data, adjusted for the full covariate set and are run once with a cohort dummy variable and again to include a cohort interaction term (*cohort). Coefficients are reported for independent and cohort interaction effects.*

*** $p<0.01$, ** $p<0.05$, * $p<0.1$

Table S7. *Linear multivariable regression models for each separate social isolation indicator to identify (1) independent effects of each social isolation indicator and (2) cohort effects in these independent effects on psychological distress, life satisfaction and self-rated general health using impute and delete data*

| All coefficients presented are for each independent social isolation indicator only (exposure) | Psychological distress<br>*Total sample N = 17,500*<br>*Males N = 8,185*<br>*Females N = 8,746*<br>coef [95% CI] | | Life satisfaction<br>*Total sample N = 18,118*<br>*Males N = 8,486*<br>*Females N = 9,008*<br>coef [95% CI] | | Self-rated general health<br>*Total sample N = 18,309*<br>*Males N = 8,600*<br>*Females N = 9,079*<br>coef [95% CI] | |
|---|---|---|---|---|---|---|
| | (1)Independent | (2)*Cohort | (1)Independent | (2)*Cohort | (1)Independent | (2)*Cohort |
| *Living alone* | | | | | | |
| *Full sample model* | .031<br>[-.017 - .079] | .023<br>[-.068 - .115] | -.207***<br>[-.257 - -.157] | .063<br>[-.034 - .161] | -.052**<br>[-.099 - -.005] | -.139***<br>[-.230 - -.048] |
| *Males only* | .033<br>[-.024 - .091] | -.030<br>[-.142 - .081] | -.190***<br>[-.255 - -.126] | .088<br>[-.035 - .210] | -.066**<br>[-.127 - -.006] | -.138**<br>[-.257 - -.020] |
| *Females only* | .046<br>[-.033 - .124] | .091<br>[-.067 - .248] | -.240***<br>[-.325 - -.156] | .026<br>[-.140 - .191] | -.044<br>[-.120 - .031] | -.123*<br>[-.268 - .022] |
| *Lack of regular contact with friends and relatives outside of the household* | | | | | | |
| *Full sample model* | .047<br>[-.050 - .145] | -.190*<br>[-.416 - .035] | -.205***<br>[-.302 - -.108] | .052<br>[-.195 - .298] | -.088*<br>[-.181 - .006] | -.070<br>[-.293 - .153] |
| *Males only* | .035<br>[-.089 - .158] | -.207<br>[-.468 - .053] | -.185***<br>[-.315 - -.054] | .004<br>[-.292 - .301] | -.086<br>[-.211 - .038] | -.061<br>[-.332 - .211] |
| *Females only* | .071<br>[-.080 - .222] | -.175<br>[-.574 - .225] | -.234***<br>[-.391 - -.077] | .096<br>[-.324 - .515] | -.073<br>[-.223 - .077] | -.100<br>[-.476 - .277] |
| *Out of employment* | | | | | | |
| *Full sample model* | .198***<br>[.154 - .243] | -.002<br>[-.083 - .079] | -.131***<br>[-.176 - -.086] | -.089**<br>[-.173 - -.005] | -.265***<br>[-.306 - -.224] | -.192***<br>[-.269 - -.114] |
| *Males only* | .307***<br>[.229 - .386] | .003<br>[-.142 - .148] | -.215***<br>[-.299 - -.131] | -.088<br>[-.239 - .064] | -.412***<br>[-.486 - -.337] | -.283***<br>[-.422 - -.144] |
| *Females only* | .158*** | -.037 | -.087*** | -.065 | -.202*** | -.139*** |

| | | | | | | |
|---|---|---|---|---|---|---|
| | [.103 - .214] | [-.140 - .066] | [-.142 - -.032] | [-.171 - .040] | [-.253 - -.152] | [-.238 - -.041] |
| *Lack of community engagement* | | | | | | |
| *Full sample model* | .012<br>[-.019 - .043] | .014<br>[-.047 - .076] | -.061***<br>[-.093 - -.029] | -.005<br>[-.072 - .062] | -.046***<br>[-.077 - -.015] | .004<br>[-.059 - .066] |
| *Males only* | -.015<br>[-.057 - .028] | .037<br>[-.050 - .124] | -.079***<br>[-.125 - -.032] | -.050<br>[-.145 - .045] | -.049**<br>[-.093 - -.004] | -.013<br>[-.103 - .078] |
| *Females only* | .038<br>[-.008 - .083] | -.005<br>[-.092 - .082] | -.045*<br>[-.091 - .001] | .026<br>[-.068 - .120] | -.044**<br>[-.087 - -.002] | .026<br>[-.057 - .110] |

Note: *All models use multiply imputed data but with cohort members deleted who had missing data on outcomes, adjusted for the full covariate set and are run once with a cohort dummy variable and again to include a cohort interaction term (*cohort). Coefficients are reported for independent and cohort interaction effects. Male and female samples do not = total sample due to missing data on the sex at birth variable.*

*** $p<0.01$, ** $p<0.05$, * $p<0.1$

Figure S1 (a, b, c and d). *Model predicted marginal mean plots for significant (p<.10) social isolation by cohort interactions on mental health outcomes*

a

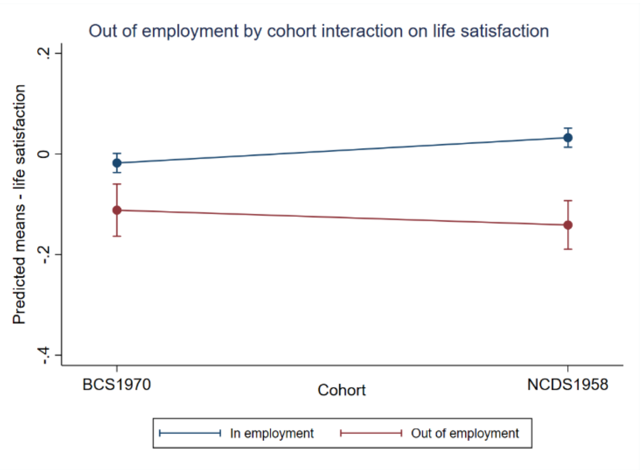

b

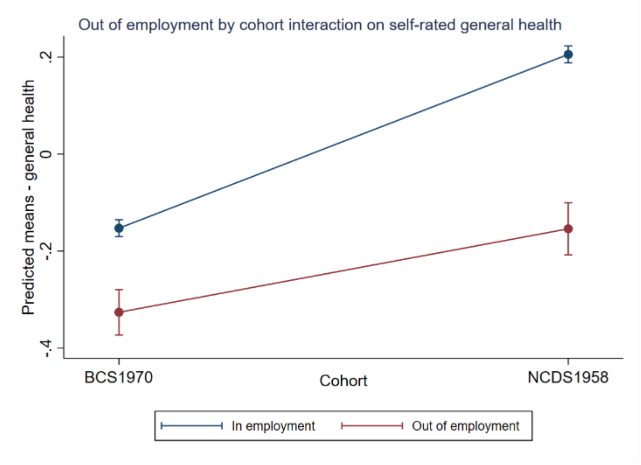

c

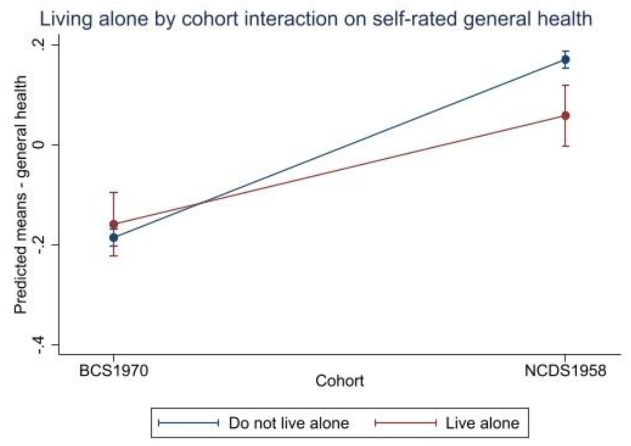

d

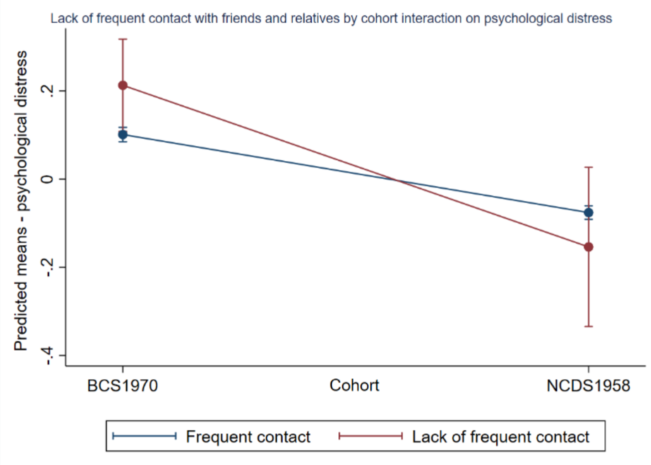

Figure S2. *Model predicted marginal mean plot for cumulative social isolation score by cohort interaction on self-rated general health*

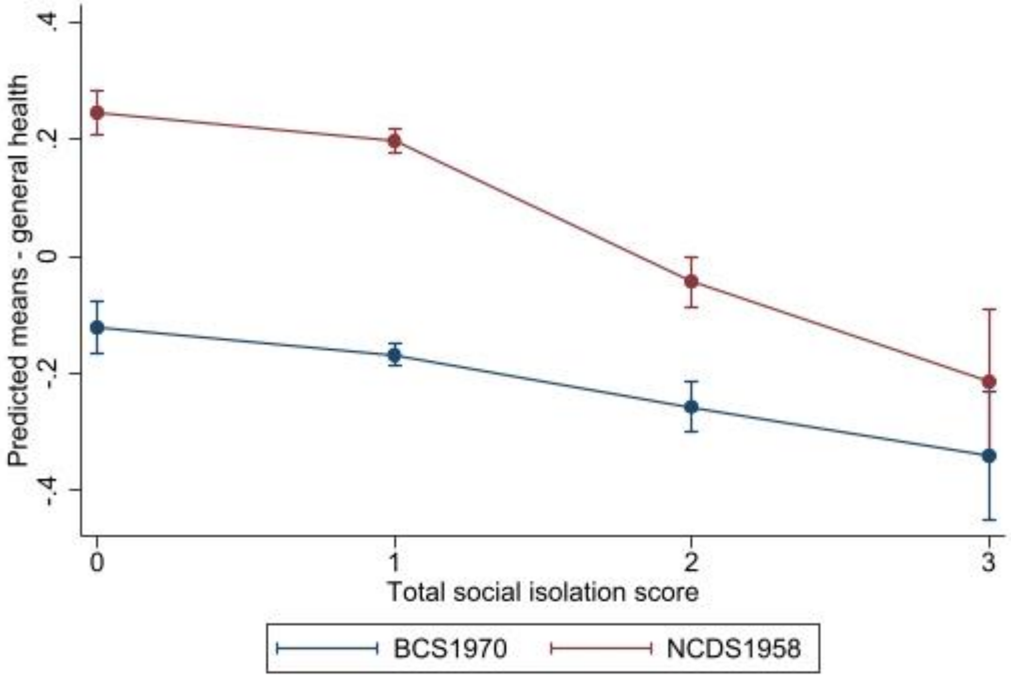

Table S8. *Linear multivariable regression models for the cumulative social isolation score to identify 1) 'dose response' effects using the cumulative social isolation score, and 2) cohort effects in any 'dose-response' associations on psychological distress, life satisfaction and self-rated general health – complete case and impute and delete models*

| All coefficients presented are for the cumulative social isolation score only (exposure) | Psychological distress coef [95% CI] *N* | | Life satisfaction coef [95% CI] *N* | | Self-rated general health coef [95% CI] *N* | |
|---|---|---|---|---|---|---|
| | (1)'Dose response' | (2)*Cohort | (1)'Dose response' | (2)*Cohort | (1)'Dose response' | (2)*Cohort |
| *Complete case* | | | | | | |
| *Full sample model* | .038*** [.012 - .064] *10,411* | -.008 [-.059 - .043] *10,411* | -.107*** [-.135 - -.080] *10,487* | -.002 [-.056 - .052] *10,487* | -.069*** [-.096 - -.042] *10,520* | -.052* [-.104 - .001] *10,520* |
| *Males only* | .019 [-.015 - .052] *5,083* | -.025 [-.092 - .042] *5,083* | -.114*** [-.151 - -.076] *5,115* | -.019 [-.094 - .056] *5,115* | -.085*** [-.123 - -.048] *5,140* | -.070* [-.144 - .004] *5,140* |
| *Females only* | .066*** [.026 - .106] *5,045* | .010 [-.069 - .089] *5,045* | -.094*** [-.134 - -.053] *5,086* | -.002 [-.082 - .079] *5,086* | -.060*** [-.099 - -.021] *5,094* | -.014 [-.091 - .063] *5,094* |
| *Impute and delete* | | | | | | |
| *Full sample model* | .065*** [.044 - .086] *17,500* | .003 [-.037 - .044] *17,500* | -.116*** [-.138 - -.093] *18,118* | -.004 [-.048 - .039] *18,118* | -.105*** [-.126 - -.084] *18,309* | -.077*** [-.117 - -.037] *18,309* |
| *Males only* | .054*** [.025 - .083] *8,185* | .002 [-.053 - .058] *8,185* | -.132*** [-.165 - -.100] *8,486* | -.006 [-.068 - .056] *8,486* | -.114*** [-.144 - -.084] *8,600* | -.091*** [-.149 - -.033] *8,600* |
| *Females only* | .082*** [.051 - .114] *8,746* | -.007 [-.066 - .053] *8,746* | -.100*** [-.133- -.067] *9,008* | .002 [-.061 - .066] *9,008* | -.100*** [-.130 - -.071] *9,079* | -.056** [-.112 - .000] *9,079* |

Note: *All models are adjusted for the full covariate set and are run once with a cohort dummy variable and again to include a cohort interaction term (*cohort). Coefficients are reported for ‘dose response’ and cohort interaction effects.*

*** $p<0.01$, ** $p<0.05$, * $p<0.1$

Table S9. *Linear multivariable regression models for the interaction between multiple social isolation indicators to identify 1) the additive and multiplicative interaction effects, and 2) cohort effects in any interaction associations on psychological distress, life satisfaction and self-rated general health using multiply imputed data m=50 (Full sample N = 32,391, Males N = 15,810, Females N = 15,221)*

| All coefficients presented are for interactions between multiple social isolation indicators (exposure) | Psychological distress coef [95% CI] | | Life satisfaction coef [95% CI] | | Self-rated general health coef [95% CI] | |
|---|---|---|---|---|---|---|
| | (1)Interaction | (2)*Cohort | (1)Interaction | (2)*Cohort | (1)Interaction | (2)*Cohort |
| *Living alone* Lack of regular contact with friends and relatives outside of the household* | | | | | | |
| *Full sample model* | .114 [-.162 - .390] | -.212 [-.815 - .391] | -.038 [-.317 - .240] | -.099 [-.744 - .546] | .018 [-.236 - .273] | -.057 [-.673 - .559] |
| *Males only* | .098 [-.245 - .442] | -.164 [-.866 - .537] | .031 [-.313 - .375] | -.372 [-1.15 - .404] | -.054 [-.374 - .267] | -.123 [-.904 - .657] |
| *Females only* | .191 [-.328 - .709] | -.402 [-1.77 - .963] | -.172 [-.711 - .368] | .454 [-.874 - 1.78] | .115 [-.348 - .577] | .195 [-.938 - 1.33] |
| *Living alone* Out of employment* | | | | | | |
| *Full sample model* | .077 [-.018 - .172] | -.111 [-.293 - .071] | -.097* [-.198 - .005] | .035 [-.160 - .231] | -.021 [-.114 - .070] | .052 [-.130 - .233] |
| *Males only* | .041 [-.089 - .171] | -.102 [-.352 - .148] | -.038 [-.175 - .099] | .033 [-.224 - .289] | .052 [-.074 - .178] | .079 [-.174 - .332] |
| *Females only* | .106 [-.058 - .270] | -.175 [-.472 - .123] | -.161* [-.337 - .015] | .052 [-.252 - .356] | -.078 [-.222 - .066] | .058 [-.228 - .345] |
| *Living alone* Lack of community engagement* | | | | | | |
| *Full sample model* | -.010 [-.107 - .086] | -.003 [-.191 - .186] | .005 [-.103 - .113] | .001 [-.216 - .217] | .003 [-.099 - .104] | -.006 [-.192 - .180] |
| *Males only* | .029 [-.104 - .161] | .056 [-.199 - .311] | -.030 [-.179 - .118] | -.050 [-.341 - .240] | .004 [-.143 - .151] | -.079 [-.331 - .173] |
| *Females only* | -.053 [-.199 - .093] | -.084 [-.404 - .237] | .052 [-.109 - .213] | .051 [-.267 - .368] | .016 [-.137 - .170] | .078 [-.227 - .383] |
| *Lack of regular contact with friends and relatives outside of the household*Out of employment* | | | | | | |

| | | | | | | |
|---|---|---|---|---|---|---|
| *Full sample model* | .030<br>[-.149 - .209] | -.007<br>[-.455 - .441] | .020<br>[-.177 - .216] | .071<br>[-.410 - .551] | .052<br>[-.113 - .218] | .077<br>[-.370 - .524] |
| *Males only* | .014<br>[-.276 - .303] | .013<br>[-.539 - .566] | .049<br>[-.255 - .354] | .091<br>[-.612 - .794] | .110<br>[-.159 - .379] | .017<br>[-.572 - .606] |
| *Females only* | .055<br>[-.214 - .323] | -.092<br>[-.778 - .593] | -.002<br>[-.287 - .283] | .056<br>[-.731 - .844] | .037<br>[-.207 - .280] | .158<br>[-.543 - .858] |
| *Lack of regular contact with friends and relatives outside of the household* Lack of community engagement* | | | | | | |
| *Full sample model* | .050<br>[-.191 - .29] | -.119<br>[-.771 - .534] | .086<br>[-.153 - .325] | -.215<br>[-.965 - .536] | -.045<br>[-.261 - .171] | .030<br>[-.656 - .717] |
| *Males only* | .098<br>[-.235 - .431] | -.116<br>[-.897 - .665] | .035<br>[-.324 - .395] | -.271<br>[-1.26 - .713] | -.046<br>[-.365 - .273] | .059<br>[-.802 - .919] |
| *Females only* | .016<br>[-.323 - .354] | -.240<br>[-1.39 - .909] | .130<br>[-.213 - .474] | -.057<br>[-1.26 - 1.15] | -.043<br>[-.362 - .275] | -.028<br>[-1.21 - 1.15] |
| *Out of employment* Lack of community engagement* | | | | | | |
| *Full sample model* | .054<br>[-.024 - .132] | .056<br>[-.100 - .212] | -.035<br>[-.117 - .047] | .012<br>[-.154 - .179] | -.070*<br>[-.151 - .011] | -.000<br>[-.149 - .149] |
| *Males only* | .036<br>[-.112 - .184] | .055<br>[-.233 - .344] | -.071<br>[-.221 - .079] | -.088<br>[-.393 - .216] | -.003<br>[-.141 - .136] | .114<br>[-.143 - .372] |
| *Females only* | .052<br>[-.055 - .159] | .056<br>[-.143 - .254] | -.012<br>[-.126 - .102] | .057<br>[-.157 - .272] | -.096**<br>[-.192- -.000] | -.071<br>[-.268 - .126] |

Note: *All models use multiply imputed data, adjusted for the full covariate set and are run once with a cohort dummy variable and again to include a cohort interaction term (*cohort). Mental health outcomes are standardised, and continuous covariates centred. Coefficients are reported for social isolation indicator interactions and cohort interaction effects. Male and female samples do not = total sample due to some missing data on the sex at birth variable.*

**** p<0.01, ** p<0.05, * p<0.1*

Table S10. *Linear multivariable regression models for the interaction between multiple social isolation indicators to identify 1) the additive and multiplicative interaction effects, and 2) cohort effects in any interaction associations on psychological distress, life satisfaction and self-rated general health using complete case data*

| All coefficients presented are for interactions between multiple social isolation indicators (exposure) | Psychological distress<br>coef [95% CI]<br>p-value<br>*N* | | Life satisfaction<br>coef [95% CI]<br>p-value<br>*N* | | Self-rated general health<br>coef [95% CI]<br>p-value<br>*N* | |
|---|---|---|---|---|---|---|
| | (1)Interaction | (2)*Cohort | (1)Interaction | (2)*Cohort | (1)Interaction | (2)*Cohort |
| *Living alone* Lack of regular contact with friends and relatives outside of the household* | | | | | | |
| *Full sample model* | .178<br>[-.289 - .644]<br>*10,929* | -.537<br>[-1.59 - .513]<br>*10,929* | -.030<br>[-.524 - .464]<br>*11,236* | -.593<br>[-1.71 - .520]<br>*11,236* | .291<br>[-.191 - .774]<br>*11,276* | -.447<br>[-1.53 - .640]<br>*11,276* |
| *Males only* | -.068<br>[-.698 - .562]<br>*5,340* | -.156<br>[-1.45 - 1.14]<br>*5,340* | -.175<br>[-.878 - .529]<br>*5,490* | -1.54**<br>[-2.99 - -.097]<br>*5,490* | .094<br>[-.609 - .796]<br>*5,518* | -.895<br>[-2.34 - .549]<br>*5,518* |
| *Females only* | .524<br>[-.302 - 1.35]<br>*5,268* | -1.33<br>[-3.25 - .590]<br>*5,268* | .140<br>[-.699 - .978]<br>*5,398* | 1.02<br>[-.923 - 2.97]<br>*5,398* | .825**<br>[.022 - 1.63]<br>*5,409* | .080<br>[-1.78 - 1.94]<br>*5,409* |
| *Living alone* Out of employment* | | | | | | |
| *Full sample model* | .046<br>[-.174 - .266]<br>*11,802* | -.206<br>[-.676 - .265]<br>*11,802* | -.128<br>[-.362 - .107]<br>*12,109* | .231<br>[-.270 - .731]<br>*12,109* | -.231***<br>[-.455 - -.007]<br>*12,155* | .319<br>[-.163 - .801]<br>*12,155* |
| *Males only* | .013<br>[-.274 - .300]<br>*5,820* | -.399<br>[-1.26 - .477]<br>*5,820* | -.099<br>[-.423 - .225]<br>*5,970* | 1.43***<br>[.447 - 2.42]<br>*5,970* | -.097<br>[-.410 - .216]<br>*5,998* | -.057<br>[-1.04 - .920]<br>*5,998* |
| *Females only* | .041<br>[-.309 - .391]<br>*5,661* | -.145<br>[-.850 - .559]<br>*5,661* | -.143<br>[-.499 - .213]<br>*5,791* | -.264<br>[-.980 - .453]<br>*5,791* | -.279<br>[-.617 - .059]<br>*5,808* | .652*<br>[-.028 - 1.33]<br>*5,808* |
| *Living alone* Lack of community engagement* | | | | | | |

| | | | | | | |
|---|---|---|---|---|---|---|
| *Full sample model* | -.082<br>[-.210 - .047]<br>*11,283* | .036<br>[-.225 - .298]<br>*11,283* | .036<br>[-.102 - .174]<br>*11,359* | -.009<br>[-.290 - .271]<br>*11,359* | .006<br>[-.127 - .139]<br>*11,398* | -.075<br>[-.346 - .196]<br>*11,398* |
| *Males only* | .018<br>[-.148 - .184]<br>*5,561* | .187<br>[-.150 - .523]<br>*5,561* | -.036<br>[-.226 - .153]<br>*5,593* | -.173<br>[-.557 - .211]<br>*5,593* | -.023<br>[-.210 - .165]<br>*5,618* | -.383**<br>[-.763 - -.003]<br>*5,618* |
| *Females only* | -.154<br>[-.358 - .051]<br>*5,439* | -.129<br>[-.555 - .297]<br>*5,439* | .122<br>[-.087 - .330]<br>*5,480* | .109<br>[-.326 - .543]<br>*5,480* | .087<br>[-.111 - .285]<br>*5,494* | .192<br>[-.221 - .605]<br>*5,494* |
| *Lack of regular contact with friends and relatives outside of the household*Out of employment* | | | | | | |
| *Full sample model* | -.271<br>[-.739 - .197]<br>*10,931* | .509<br>[-.694 - 1.71]<br>*10,931* | .138<br>[-.359 - .635]<br>*11,238* | .123<br>[-1.15 - 1.40]<br>*11,238* | .473*<br>[-.011 - .956]<br>*11,278* | -.854<br>[-2.10 - .389]<br>*11,278* |
| *Males only* | .211<br>[-.496 - .917]<br>*5,343* | -.354<br>[-1.78 - 1.07]<br>*5,343* | .269<br>[-.523 - 1.06]<br>*5,493* | .180<br>[-1.41 - 1.77]<br>*5,493* | .410<br>[-.377 - 1.20]<br>*5,521* | -.915<br>[-2.50 - .669]<br>*5,521* |
| *Females only* | -.320<br>[-1.01 - .366]<br>*5,267* | - | .159<br>[-.539 - .857]<br>*5,397* | - | .547<br>[-.118 - 1.21]<br>*5,408* | - |
| *Lack of regular contact with friends and relatives outside of the household* Lack of community engagement* | | | | | | |
| *Full sample model* | .197<br>[-.137 - .531]<br>*10,419* | -.627<br>[-1.76 - .505]<br>*10,419* | .312*<br>[-.043 - .667]<br>*10,495* | .097<br>[-1.11 - 1.30]<br>*10,495* | -.214<br>[-.559 - .132]<br>*10,528* | .713<br>[-.461 - 1.89]<br>*10,528* |
| *Males only* | .319<br>[-.100 - .739]<br>*5,088* | -.446<br>[-1.90 - 1.00]<br>*5,088* | .310<br>[-.163 - .782]<br>*5,120* | .079<br>[-1.56 - 1.72]<br>*5,120* | -.130<br>[-.599 - .340]<br>*5,145* | .938<br>[-.690 - 2.57]<br>*5,145* |
| *Females only* | .078<br>[-.450 - .606]<br>*5,048* | -.846<br>[-2.59 - .901]<br>*5,048* | .355<br>[-.181 - .891]<br>*5,089* | -.026<br>[-1.80 - 1.75]<br>*5,089* | -.284<br>[-.797 - .230]<br>*5,097* | .420<br>[-1.28 - 2.12]<br>*5,097* |

| *Out of employment* Lack of community engagement* | | | | | | |
|---|---|---|---|---|---|---|
| *Full sample model* | .197** [.042 - .353] *11,285* | -.096 [-.417 - .225] *11,285* | -.179** [-.346 - -.012] *11,361* | .002 [-.342 - .345] *11,361* | -.206** [-.368 - -.044] *11,400* | -.209 [-.542 - .124] *11,400* |
| *Males only* | .186 [-.104 - .475] *5,564* | -.526* [-1.13 - .075] *5,564* | -.381** [-.711- -.051] *5,596* | .139 [-.547 - .824] *5,596* | -.129 [-.455 - .196] *5,621* | .421 [-.256 - 1.10] *5,621* |
| *Females only* | .199** [.000 - .398] *5,438* | .003 [-.409 - .414] *5,438* | -.127 [-.330 - .076] *5,479* | -.023 [-.443 - .397] *5,479* | -.218** [-.411 - -.025] *5,493* | -.400** [-.799 - -.001] *5,493* |

Note: *All models use complete case data, adjusted for the full covariate set and are run once with a cohort dummy variable and where significant, again to include a cohort interaction term (*cohort) and for males and females only. Mental health outcomes are standardised, and continuous covariates centred. Coefficients are reported for social isolation indicator interactions and cohort interaction effects. Male and female samples do not = total sample due to some missing data on the sex at birth variable. - = no observations in the sample.*

**** p<0.01, ** p<0.05, * p<0.1*

Table S11. *Linear multivariable regression models for the interaction between multiple social isolation indicators to identify 1) the additive and multiplicative interaction effects, and 2) cohort effects in any interaction associations on psychological distress, life satisfaction and self-rated general health using impute and delete data*

| All coefficients presented are for interactions between multiple social isolation indicators (exposure) | Psychological distress<br>*Total sample N = 17,500*<br>*Males N = 8,185*<br>*Females N = 8,746*<br>coef [95% CI]<br>p-value | | Life satisfaction<br>*Total sample N = 18,118*<br>*Males N = 8,486*<br>*Females N = 9,008*<br>coef [95% CI]<br>p-value | | Self-rated general health<br>*Total sample N = 18,309*<br>*Males N = 8,600*<br>*Females N = 9,079*<br>coef [95% CI]<br>p-value | |
|---|---|---|---|---|---|---|
| | (1)Interaction | (2)*Cohort | (1)Interaction | (2)*Cohort | (1)Interaction | (2)*Cohort |
| *Living alone* Lack of regular contact with friends and relatives outside of the household* | | | | | | |
| *Full sample model* | .238<br>[-.123 - .600] | -.490<br>[-1.33 - .351] | -.044<br>[-.409 - .320] | -.231<br>[-1.13 - .666] | .064<br>[-.269 - .397] | -.197<br>[-1.07 - .672] |
| *Males only* | .154<br>[-.320 - .628] | -.336<br>[-1.35 - .675] | .145<br>[-.351 - .642] | -.907<br>[-2.08 - .262] | -.050<br>[-.525 - .425] | -.371<br>[-1.44 - .693] |
| *Females only* | .357<br>[-.298 - 1.01] | -.848<br>[-2.62 - .923] | -.359<br>[-1.06 - .338] | .985<br>[-.930 - 2.90] | .211<br>[-.332 - .755] | .365<br>[-1.25 - 1.98] |
| *Living alone* Out of employment* | | | | | | |
| *Full sample model* | .165**<br>[.033 - .297] | -.288**<br>[-.544 - -.032] | -.197***<br>[-.335 - -.058] | .131<br>[-.146 - .408] | -.049<br>[-.169 - .072] | .123<br>[-.114 - .359] |
| *Males only* | .082<br>[-.080 - .244] | -.311*<br>[-.641 - .019] | -.024<br>[-.206 - .157] | .107<br>[-.272 - .486] | .137<br>[-.027 - .300] | .146<br>[-.178 - .471] |
| *Females only* | .223**<br>[.009 - .437] | -.406*<br>[-.826 - .014] | -.333***<br>[-.556 - -.111] | .137<br>[-.309 - .583] | -.137<br>[-.324 - .051] | .174<br>[-.196 - .544] |
| *Living alone* Lack of community engagement* | | | | | | |
| *Full sample model* | -.022<br>[-.140 - .095] | .023<br>[-.218 - .264] | -.008<br>[-.130 - .113] | .016<br>[-.244 - .277] | .013<br>[-.105 - .130] | -.041<br>[-.275 - .194] |
| *Males only* | .058<br>[-.096 - .213] | .104<br>[-.223 - .431] | -.081<br>[-.251 - .089] | -.066<br>[-.421 - .288] | .004<br>[-.157 - .165] | -.174<br>[-.504 - .157] |

| | | | | | | |
|---|---|---|---|---|---|---|
| *Females only* | -.084 [-.269 - .100] | -.088 [-.473 - .297] | .078 [-.115 - .271] | .070 [-.323 - .462] | .055 [-.127 - .238] | .104 [-.259 - .466] |
| *Lack of regular contact with friends and relatives outside of the household*Out of employment* | | | | | | |
| *Full sample model* | .024 [-.251 - .299] | .047 [-.603 - .697] | .061 [-.221 - .343] | .114 [-.544 - .773] | .094 [-.146 - .333] | .055 [-.550 - .660] |
| *Males only* | -.007 [-.432 - .419] | .146 [-.716 - 1.01] | .163 [-.329 - .656] | .154 [-.841 - 1.15] | .300 [-.104 - .703] | -.102 [-.925 - .722] |
| *Females only* | .042 [-.358 - .442] | -.156 [-1.20 - .888] | -.017 [-.426 - .391] | .107 [-.938 - 1.15] | .015 [-.326 - .355] | .200 [-.763 - 1.16] |
| *Lack of regular contact with friends and relatives outside of the household* Lack of community engagement* | | | | | | |
| *Full sample model* | .127 [-.158 - .413] | -.253 [-1.07 - .561] | .133 [-.174 - .440] | -.312 [-1.21 - .588] | -.056 [-.333 - .221] | .134 [-.708 - .977] |
| *Males only* | .232 [-.153 - .618] | -.308 [-1.28 - .666] | .048 [-.370 - .467] | -.413 [-1.53 - .708] | -.060 [-.449 - .328] | .186 [-.853 - 1.23] |
| *Females only* | .057 [-.358 - .473] | -.488 [-2.05 - 1.08] | .195 [-.250 - .641] | .075 [-1.55 - 1.70] | -.044 [-.472 - .385] | .038 [-1.51 - 1.59] |
| *Out of employment* Lack of community engagement* | | | | | | |
| *Full sample model* | .109** [.010 - .208] | .060 [-.122 - .242] | -.037 [-.140 - .067] | .027 [-.166 - .221 | -.127*** [-.217 - -.036] | .042 [-.142 - .225] |
| *Males only* | .103 [-.087 - .294] | .101 [-.269 - .470] | -.140 [.358 - .078] | -.226 [-.639 - .187] | .013 [-.183 - .209] | .282 [-.103 - .666] |
| *Females only* | .079 [-.042 - .201] | .051 [-.182 - .284] | .005 [-.121 - .131] | .100 [-.144 - .344] | -.144*** [-.251 - -.036] | -.059 [-.286 - .168] |

Note: *All models use impute and delete data, adjusted for the full covariate set and are run once with a cohort dummy variable and where significant, again to include a cohort interaction term (*cohort) and for males and females only. Mental health outcomes are standardised, and continuous covariates centred. Coefficients are reported for social isolation indicator interactions and cohort interaction effects. Male and female samples do not = total sample due to some missing data on the sex at birth variable.*

**** p<0.01, ** p<0.05, * p<0.1*

*Latent Class Analysis Methodology*

Latent class analysis was adopted to identify groups of individuals showing qualitatively similar patterns in their social conditions (Weller et al., 2020). First, latent class analysis generates probabilities for who belongs to which group or class. Then, the models describe the relationship between classes and the observed variables. To understand the optimal number of social isolation groups within the current data, models were run estimating an increasing number of classes between 1 and 5. When selecting the class solution, goodness-of-fit statistics were used in combination with a theoretical interpretation (Weller et al., 2020). The Bayesian information criteria (BIC) is the most commonly used criteria, with a lower BIC indicating better fit (Killian et al., 2019). We assessed both the BIC and the adjusted Bayesian information criteria (ABIC). The Akaike information criterion (AIC) indicated the adequacy of different non-nested models, where lower IC indicates a more adequate model (Wagenmakers & Farrell, 2004). In addition to these ICs, we calculated the entropy and posterior class-membership probabilities (Celeux & Soromenho, 1996) where an entropy value closer to 1 is preferable, and average latent posterior probabilities of greater than .90 are desirable (Muthén & Muthén, 2000). The Vuong-Lo-Mendell-Rubin adjusted likelihood ratio test also helped determine the final class solution (Lo et al., 2001; Vuong et al., 1989). Finally, we considered the general rule that classes should not contain less than 5% of the sample (Shanahan et al., 2013). Latent classes were assessed by sex and cohort. A Full Information Maximum Likelihood (FIML) approach to handling missing data was applied in Mplus eighth edition (Muthén & Muthén, 2017) for the latent class analyses.

Table S12. *Results from the latent class analyses (1-5 classes) including goodness-of-fit statistics (full sample with 1970 BCS and 1958 cohorts combined N=19,902)*

| Class solution | BIC | ABIC | AIC | Entropy | Average latent posterior probabilities (>.90) yes/no | Vuong-Lo-Mendell-Rubin adjusted likelihood ratio test (p-value) | Any classes with less than 5% of the sample yes/no |
|---|---|---|---|---|---|---|---|
| *1* | 48779.359 | 48766.647 | 48747.764 | - | - | - | - |
| *2* | 48719.095 | 48690.493 | 48648.008 | .985 | yes | .000 | no |
| *3* | 48732.183 | 48687.692 | 48621.603 | .932 | no | .000 | yes |
| *4* | 48776.514 | 48716.133 | 48626.441 | .836 | no | .108 | yes |
| *5* | 48823.570 | 48747.299 | 48634.004 | .462 | no | .732 | yes |

Note: *BIC = Bayesian information criteria, ABIC = adjusted Bayesian information criteria, AIC = Akaike information criterion. Sample based on FIML approach in Mplus*

Given similar BIC, ABIC, AIC and entropy values when compared to the two-class solution, and a significant Vuong-Lo-Mendell-Rubin adjusted likelihood ratio test indicating a better fit to the data, the three-class solution was identified as the optimal class solution for these data. Despite all groups in the three-class solution being characterised by a high probability of limited community engagement, the first class included those who also lived alone, the second class those who lacked regular contact with friends and relatives outside the household, and the third class had a low probability of isolation across all contexts except the community.

Table S13. *Class membership (N(%)) for full sample and by sex and cohort*

| Class | Full sample *N=19,902* | Males *N= 8,371* | Females *N=9,083* | 1970 BCS *N=9,802* | 1958 NCDS *N=10,100* |
|---|---|---|---|---|---|
| *1 – living alone and lacking community engagement* | *393*(1.97) | *156*(1.86) | *111*(1.22) | *223*(2.28) | *170*(1.68) |
| *2 – lack of frequent contact with friends and relatives outside the household and lacking community engagement* | *392*(1.97) | *170*(2.03) | *124*(1.37) | *317*(3.23) | *75*(0.74) |
| *3 – low isolation except lack of community engagement* | 19,117(96.06) | *8,045*(96.11) | *8,848*(97.41) | *9,262*(94.49) | *9,855*(97.57) |

Note: *sample based on FIML approach in Mplus and complete case sex*
